# Estimating the contribution of transmission in primary healthcare clinics to community-wide TB disease incidence, and the impact of infection prevention and control interventions, in KwaZulu-Natal, South Africa

**DOI:** 10.1101/2021.08.02.21260844

**Authors:** Nicky McCreesh, Aaron S Karat, Indira Govender, Kathy Baisley, Karin Diaconu, Tom A Yates, Rein MGJ Houben, Karina Kielmann, Alison D Grant, Richard G White

## Abstract

**Background:** There is a high risk of *Mycobacterium tuberculosis* (*Mtb*) transmission in healthcare facilities in high burden settings. Recent World Health Organization guidelines on tuberculosis infection prevention and control (IPC) recommend a range of measures to reduce transmission in healthcare and institutional settings. These were evaluated primarily based on evidence for their effects on transmission to healthcare workers in hospitals. To estimate the overall impact of IPC interventions, it is necessary to also consider their impact on community-wide tuberculosis incidence and mortality.

**Methods:** We developed an individual-based model of *Mtb* transmission between household members, in primary healthcare clinics (PHCs), and in other congregate settings; drug sensitive and multidrug resistant tuberculosis disease development and resolution; and HIV and antiretroviral therapy (ART) and their effects on tuberculosis. The model was parameterised using data from a high HIV prevalence, rural/peri-urban community in KwaZulu-Natal, South Africa, including data on social contact in clinics and other settings by sex, age group, and HIV/ART status; and data on the prevalence of tuberculosis in clinic attendees and the general population. We estimated the proportion of disease in adults that resulted from transmission in PHC clinics in 2019, and the impact of a range of IPC interventions in clinics on community-wide TB incidence and mortality.

**Results:** We estimate that 7.6% (plausible range 3.9-13.9%) of drug sensitive and multidrug resistant tuberculosis in adults resulted from transmission in PHC clinics in the study community in 2019. The proportion is higher in HIV-positive people, at 9.3% (4.8%-16.8%), compared to 5.3% (2.7%-10.1%) in HIV-negative people. We estimate that IPC interventions could reduce the number of incident TB cases in the community in 2021-2030 by 3.4-8.0%, and the number of deaths by 3.0-7.2%.

**Conclusions:** A non-trivial proportion of tuberculosis results from transmission in PHC clinics in the study communities, particularly in HIV-positive people. Implementing IPC interventions could lead to moderate reductions in disease burden. We therefore recommend that IPC measures in clinics should be implemented both for their benefits to staff and patients, but also for their likely effects on TB incidence and mortality in the surrounding community.

## Introduction

Tuberculosis (TB) is a major global public health problem, killing an estimated 1.4 million people in 2019^1^. There is a high risk of transmission in healthcare facilities in high TB burden settings, evidenced by the elevated rate of tuberculosis in healthcare workers^2^. Updated World Health Organization guidelines on TB infection prevention and control (IPC) recommend a wide range of measures to reduce transmission in healthcare and institutional settings, ranging from triaging people with TB symptoms to installing ultraviolet germicidal irradiation systems (UVGI)^2^. These measures were evaluated and implemented as recommendations in the guidelines primarily based on evidence on their effects on risk to healthcare workers, and in hospitals settings.

Protecting healthcare workers should be a key concern of TB control programmes. However, the motivation for, and potential benefits of, IPC interventions in clinics extend beyond the reductions in disease burden among clinic staff. While healthcare workers and other clinic staff are at the highest risk of infection in clinics, due to their longer durations of exposure, the numbers of patients and other clinic attendees are far higher than numbers of staff. It is therefore likely that a large proportion of clinic-acquired TB is in patients and other clinic attendees. As a consequence, it is imperative that the impact on TB incidence in the wider community is considered when estimating the likely impacts of IPC measures.

Estimating the contribution of transmission in clinics (or other congregate settings) to overall community-wide disease burden is challenging. Taylor *et al*. used data on ventilation rates and a Wells-Riley approach to estimate a 0.03% risk of infection to patients per clinic visit. This approach is heavily dependent on estimates of mean quanta production rates, however, about which there is considerable uncertainty (their sensitivity analysis gave a wide range of 0.02-0.35%). Andrews *et al* also used a Wells-Riley based approach to determine infection risk by location (although not clinics), but removed the dependence on an assigned value for the quanta production rate by using data on contact time in multiple types of location, and calibrating their model to the prevalence of infection by age^3^.

In this work, we used a social contact data-based approach similar to that adopted by Andrews *et al*., but used an individual-based model that includes HIV/antiretroviral therapy (ART) and TB disease development and resolution, and calibrated the model to overall disease incidence. This allowed us to determine the contribution of primary healthcare (PHC) clinics not only to the incidence of infection, but also to community-wide disease incidence and mortality. This is important for determining the true contribution of clinics-based transmission to disease burden, due to the increased rates of clinic attendance by people at increased risk of progression to disease^4^. We also incorporated empirical data on the increased prevalence of TB in PHC clinic attendees compared to the general population, something that acts to amplify transmission in clinics^4^.

The IPC interventions we simulated were identified and parameterised through a rigorous multi-disciplinary approach. This work forms part of the *Umoya omuhle* project, that used a whole systems approach to study IPC in primary healthcare facilities in South Africa. As part of the project, system dynamics modelling was used to identify potential IPC interventions that local policy makers and health professionals active at clinic and province levels ranked highly in terms of both feasibility of implementation and perceived likely impact on overall and MDR *Mtb* transmission in clinics^5^.

## Methods

### Social contact data

#### Data collection

A social contact survey was conducted in the catchment areas of two primary healthcare clinics in the southern section of the Africa Health Research Institute (AHRI) demographic surveillance area (DSA)^6^, in March-December 2019. 3093 adults (aged 18 years and over) were sampled, stratified by local area.

Respondents were asked to list all indoor locations visited and transport used on an assigned day in the week before the survey. For each location visited (including their own home) and transport used, they were asked for further details, including the type of location, the duration of time they spent there, and the number of other people present. Respondents were also asked the number of times they had visited clinics in the six months before the interview, and how long they spent at the clinic and a cross-sectional estimate of the number of people present on their last visit.

Further details of the social contact survey are given in the supporting material and in McCreesh *et al* 2021^7^. Ethical approval was granted by the Biomedical Research Ethics Committee (BREC) of the University of KwaZulu-Natal (BE662/17) and the London School of Hygiene & Tropical Medicine (14640).

#### Analysis

For each location visited on the assigned day, adult contact times were calculated as the reported number of adults present (capped at a maximum of 100) multiplied by the duration of time spent at the location. Respondents who reported being HIV positive were considered to be HIV positive. Otherwise, respondents were considered to be HIV negative/unknown.

#### Transmission model

We developed an individual-based model (IBM) of social contact behaviour, *Mtb* transmission by location, TB disease development and treatment, and HIV and ART. Simulated individuals were created at age 15 years, and died at age 80 years, with additional TB, HIV and background mortality occurring between those ages. People aged <15 years were not simulated, as the risk of transmission from children is low^8^, and contact data were not available from children from the study population.

Individuals could be uninfected with *Mtb*, have a latent infection, have smear-negative disease, have smear-positive disease, or be on treatment for TB. Drug sensitivity was represented as drug sensitive (DS-TB) or multidrug resistant (MDR-TB). HIV was also simulated, and individuals could be HIV-negative; HIV-positive, not on ART; or HIV-positive, on ART.

Each simulated individual was a member of a household, with the household size distribution taken from empirical data^6^. Each individual had the same amount of contact time with each household member each month. Mean clinic contact time per month in the model varied by sex and HIV/ART status strata, and within those strata, by whether someone was assigned to a high or low clinic visiting group. Mean clinic contact time could also be higher in people with untreated TB disease, to allow the model to be fitted to empirical data on the prevalence of TB in clinic attendees relative to the general population^9^. Finally, contact time occurring in all other indoor locations (including transport) was simulated, varying by sex, age group, and HIV/ART status. Contact time was parameterised using data from the social contact survey.

A baseline rate of transmission per minute contact between each person with untreated TB and each person uninfected or latent person was simulated. This was adjusted according to the smear status of the person with TB; whether the person at risk was uninfected or had a latent infection (giving partial protection against reinfection), and, if latent, by their HIV/ART status; and the assumed mean rate of ventilation in locations of that type (household, clinic, or other).

The model was fitted by hand to empirical data from the social contact study population and from KwaZulu-Natal province.

A full description of the model and parameters is given in the supporting material.

#### Interventions

Seven potential IPC interventions had been identified in qualitative research and system dynamics modelling exercises conducted as part of the *Umoya omuhle* project^5^. The effect of the interventions on patient contacts and infection risk in clinics were estimated in previous modelling work, using a within-clinics model that simulated the flow of patients through clinics, and ventilation rates and infection risk in clinic waiting areas^10^. The interventions were:

1. **Windows & doors**. Ensuring windows and doors in waiting areas are kept open at all times. This was estimated to reduce the rate of transmission to clinic attendees by 55% (interquartile range [IQR] 25-72%).
2. **Retrofits**. Building retrofits are changes to the building to improve ventilation rates. This could include installing lattice brickwork or whirlybird fans. Due to the large amount of variation between clinic spaces in the types of building retrofits that would be suitable, and the lack of sufficient data on the effects of the retrofits on ventilation rates in different types of spaces, we did not model specific retrofits or packages of retrofits. Instead, in the within clinics model, we simulated an undefined package of retrofits that are sufficient to increase air changes per hour to a minimum of 12 in all waiting rooms, chosen in line with WHO guidelines^2 11^. This was estimated to reduce the rate of transmission to clinic attendees by 45% (IQR 16-64%)
3. **Ultraviolet Germicidal Irradiation (UVGI) system**. We assumed in this intervention that appropriate and well maintained UVGI systems are installed in all indoor clinic waiting areas. This was estimated to reduce the rate of transmission to clinic attendees by 77% (IQR 64-85%)
4. **Masks**. We simulated a scenario where 70% of patients wear surgical masks 90% of the time. This was estimated to reduce the rate of transmission to clinic attendees by 47% (IQR 42-50%)
5. **CCMDD coverage**. South Africa’s Central Chronic Medicine Dispensing and Distribution (CCMDD) programme is designed to allow patients with stable chronic health conditions to collect their medicines from convenient locations, such as local pharmacies^12^. This means that they do not need to queue at clinics unnecessarily. The purpose of this intervention was to increase the utilisation of CCMDD and similar programmes by eligible patients, and to ensure that pick-up points do not require patients to queue at clinics. This was estimated to reduce mean clinic contact time per visit by 28% (IQR 9-42%) for patients on ART and 13% (8-19%) for all other patients, reducing the overall rate of transmission to clinic attendees by 22% (IQR 12-32%).
6. **Queue management system with outdoor waiting area**. This intervention combined a large, well ventilated, covered outdoor waiting area with a queue management system. This was estimated to reduce the rate of transmission to clinic attendees by 83% (IQR 76-88%)
7. **Appointment system**. In this intervention, we simulated a date-time appointment system to reduce clinic overcrowding, through spacing out the arrival times of patients. This was estimated to reduce the overall rate of transmission to clinic attendees by 62% (IQR 45-75%)

The estimated effects of the interventions on patient contacts and infection risk in clinics from the within-clinics model were used to parameterise the effects of the interventions on contact rates and transmission probabilities in clinics in this model, allowing their wider effects on community-level disease incidence to be estimated. The interventions were implemented in the model from 2021. Full details are given in the supporting material.

#### Uncertainty analyses

A number of univariate sensitivity analyses were conducted, exploring the effects of uncertainty in clinic contact time, the proportion of disease from transmission between household members, ventilation rates in clinics, the prevalence of TB in clinic attendees relative to the general population, the rate at which people switch between the high and low clinic visiting groups, clinic visiting rates in HIV-people who are not on ART, and future HIV incidence. These sensitivity analyses were used to construct a plausible range around the estimated proportion of disease that results from transmission in clinics, and estimated intervention impact. Full details are given in the supporting material.

#### Proportion of disease from transmission in clinics that is in clinic staff

In the mathematical model, we only consider transmission to adult clinic attendees. Clinic staff are also at risk of infection in clinics however. We used a simple method to obtain a rough estimate of the proportion of disease that results from transmission in clinics that is in clinic staff, assuming that all clinic staff who are at elevated risk of infection from transmission in clinics have the same exposure to TB outside the clinic as the general population, and that all excess TB in clinic staff results from transmission in clinics. Full details are given in the supporting material.

## Results

### Social contact data

A total of 1704 individuals were interviewed. A description of respondent characteristics and reported contact time is given in the supporting material.

### Fit to data

The model fit well to all the fitting targets, in the main scenario and the sensitivity analyses scenarios (Figure 1 and supporting material Table S12).

**Figure 1.**
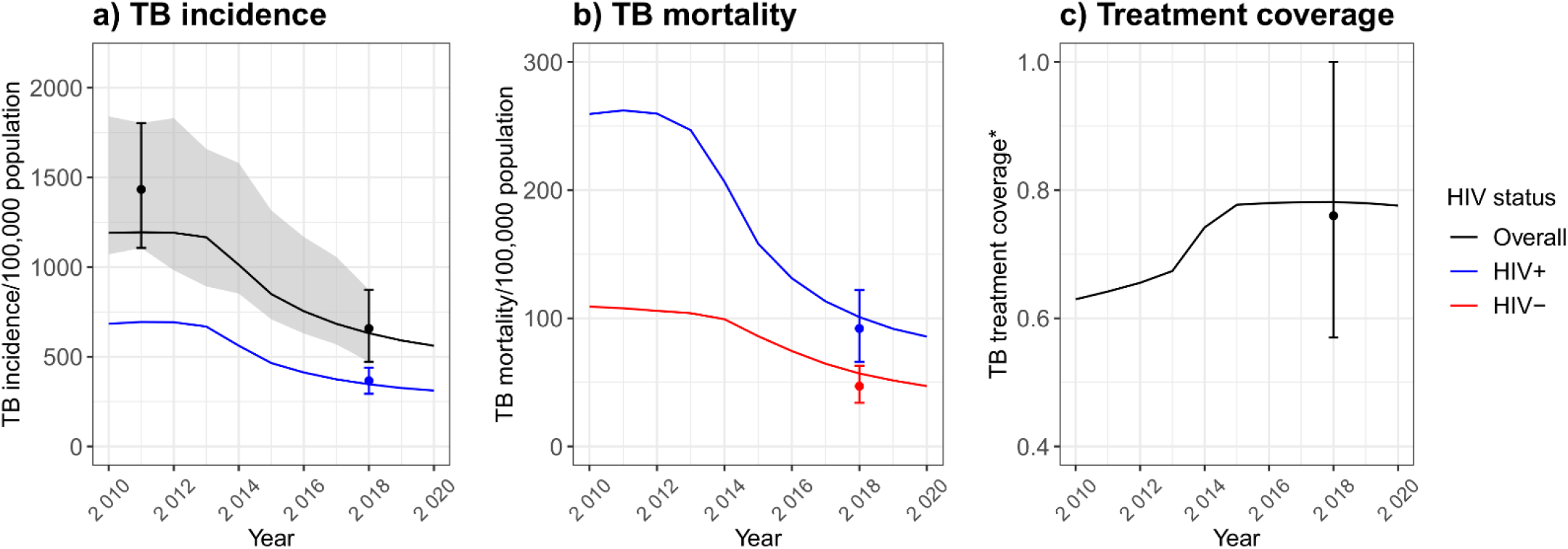
Model fit to estimated TB incidence, mortality, and treatment coverage. *Treatment coverage is calculated as the ratio of the number of people starting treatment in a year to the estimated number of people developing TB in the same year. Lines over time show model output. Points and error bars show the fitting targets, based on empirical data. The ribbon in plot a) shows the empirical estimates over time. Empirical estimates over time were not available for KwaZulu-Natal for the other fitting outputs shown here.

**Figure 2.**
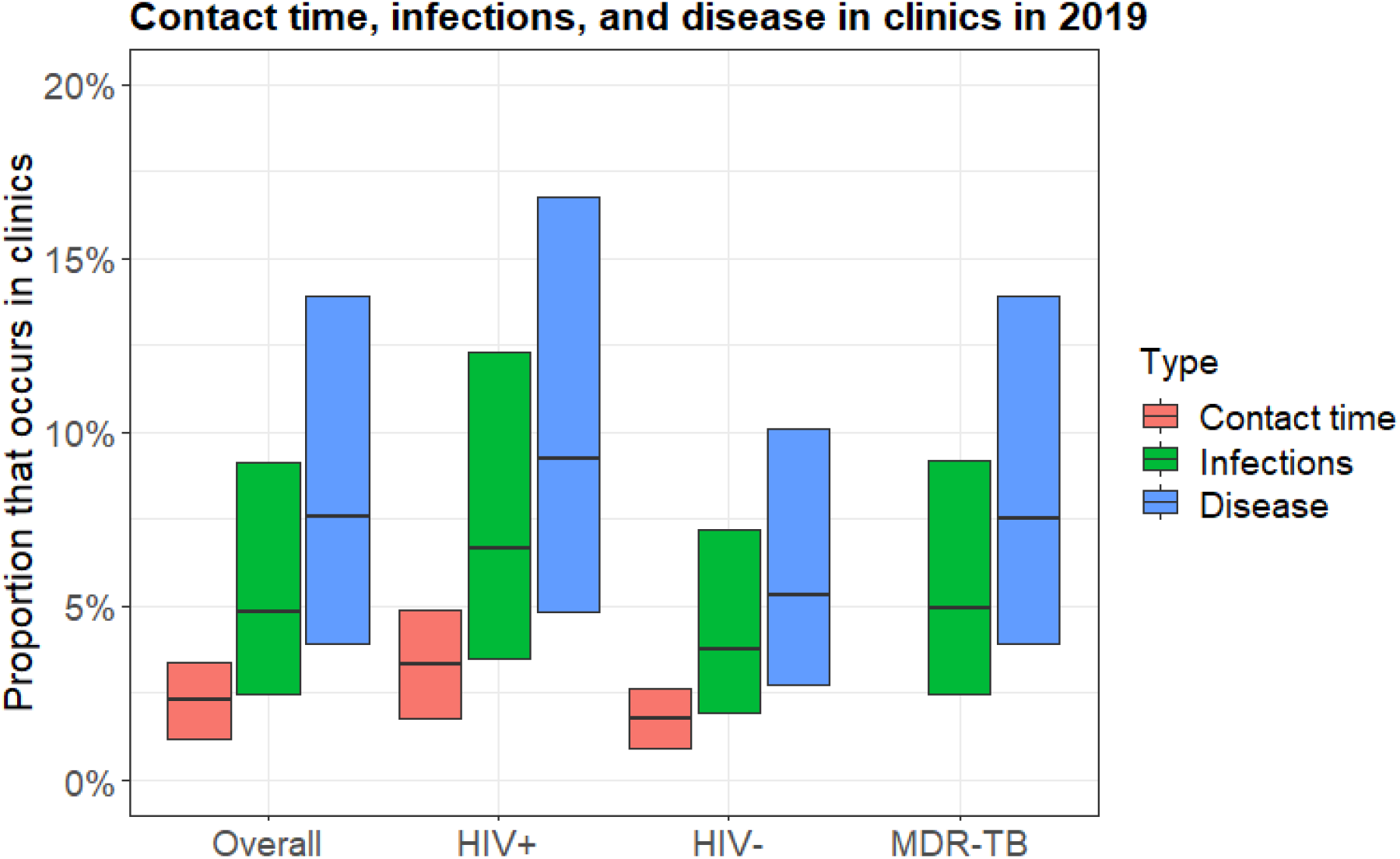
The estimated proportion of contact time and infections that occurs in clinics, and the proportion of disease that resulted from transmission in clinics in the study population in 2019, overall, in HIV-positive people, HIV-negative people, and for MDR-TB. For contact time, the central bar shows the proportion for the best estimate scenario, and the range of the bars shows the proportions in the contact time in clinics sensitivity analysis. For infections and disease, the central horizontal bar shows the best estimate, and the range of the bars shows the most extreme results from the sensitivity analyses.

### Proportion of disease from transmission in clinics

Overall, we estimate that 2.3% (plausible range 1.2-3.4%) of contact time by adults occurred in clinics in the study communities in 2019, leading to 4.9% (2.5-9.1%) of overall and MDR infections, and 7.6% (3.9-13.9%) of overall and MDR disease. The proportion of all TB disease that resulted from transmission in clinics was higher in HIV-positive people, at 9.3% (range 4.8%-16.8%), and lower in HIV-negative people, at 5.3% (2.7%-10.1%).

### Intervention impact

Opening windows and doors reduced the total number of incident TB cases in the community in 2021-2030 by 5.3% (range 1.3-12.5%), simple clinic retrofits by 4.3% (0.8-11.2%), UVGI systems by 7.4% (3.2-14.7%), surgical mask wearing by patients by 4.5% (2.1-8.8%), increased CCMDD coverage by 3.4% (0.7-8.7%), queue management systems with outdoor waiting areas by 8.0% (3.8-15.2%), and appointment systems by 5.9% (2.2-12.9%) (Figure 3). Reductions in MDR-TB cases were similar to reductions in all TB cases (Supporting figure S6).

**Figure 3.**
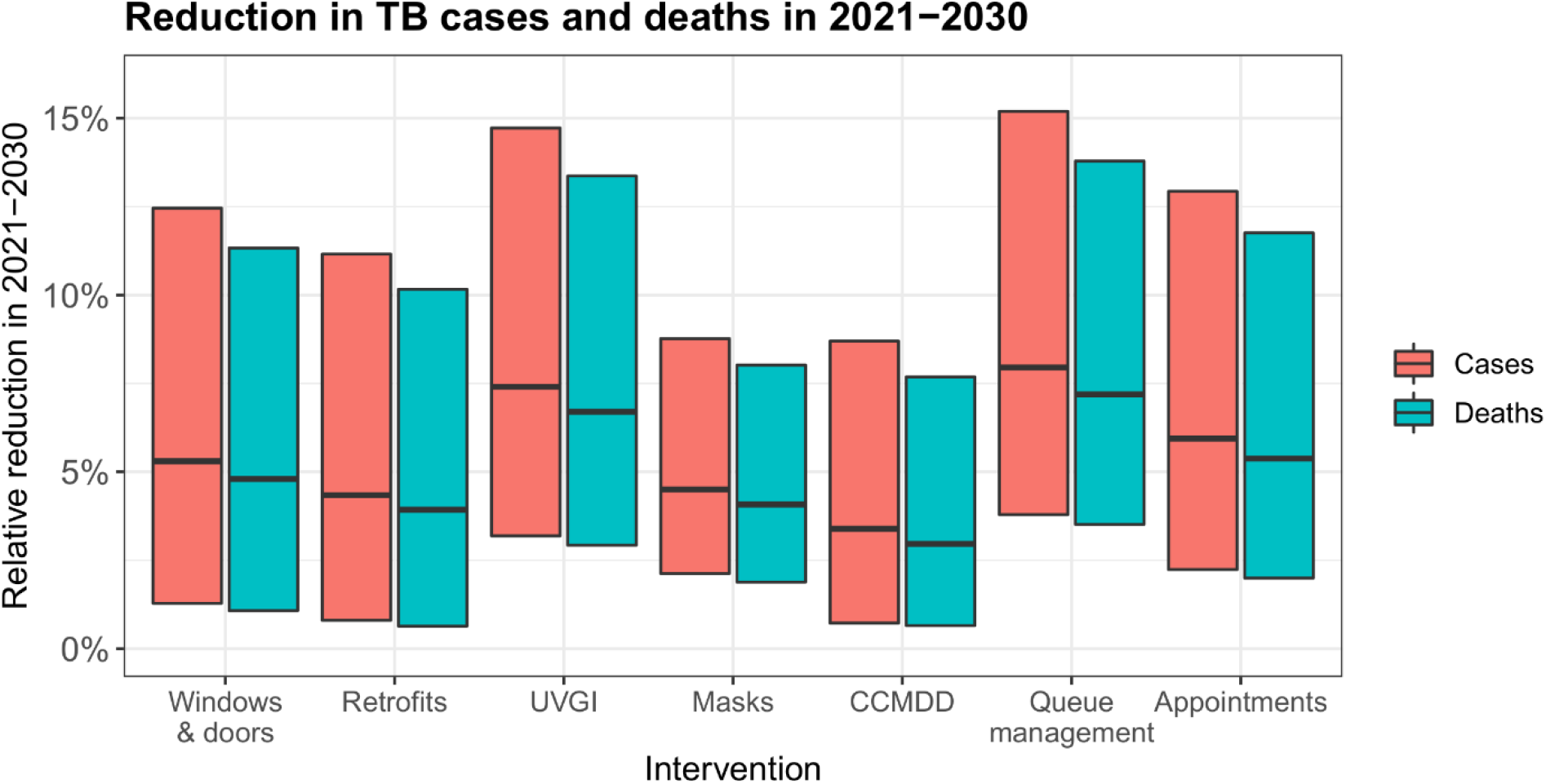
The estimated reduction in TB cases and deaths in the study population in 2021-2030 resulting from the proposed infection control interventions. The central horizontal bar shows the best estimate, and the range of the bars shows the most extreme results from the sensitivity analyses.

Reductions in TB deaths were 9.5-12.6% lower than reductions in cases, reflecting the time lag between developing disease and dying from TB. The reductions in deaths ranged from 3.0% (range 0.7-10.1%) for increased CCMDD coverage, to 7.2% (range 2.7-13.8%) for queue management systems with outdoor waiting areas.

### Proportion of disease from transmission in clinics that is in clinic staff

We estimate that in the study community, an average of 7.1% (95% plausible range 2.3-16.7%) of all disease in adults resulting from transmission in clinics occurs in clinic staff.

## Discussion

In this paper, we estimate that 7.6% (plausible range 3.9-13.9%) of tuberculosis in adults results from transmission in PHC clinics in a high HIV prevalence, rural/peri-urban setting in KwaZulu-Natal, South Africa. The proportion is higher in HIV-positive people, at an estimated 9.3% (range 4.8%-16.8%), compared to 5.3% (2.7%-10.1%) in HIV-negative people. We estimate that IPC interventions in PHC clinics could reduce the number of incident TB cases in the community in 2021-2030 by 3.4-8.0%, and deaths by 3.0-7.2%. These findings further strengthen the case for an increased emphasis on IPC in clinics, not just as a tool for protecting clinic staff and patients, but as method for reducing community-wide TB incidence and mortality.

Our findings highlight the importance of considering contact saturation^13^ and the population at risk when estimating the proportion of disease that results from transmission in different types of setting. We estimate that 4.9% of infections occur in clinics, more than double the estimated 2.3% of contact time occurring in clinics. This reflects the fact that contacts between household members are repeated, reducing their overall importance to transmission, and increasing the importance of other settings. It also reflects higher rates of clinic attendance by people with potentially infectious TB. We estimate that an even higher proportion of disease results from transmission in PHC clinics, at 7.7%. This reflects both the additional effects of contact saturation and repeated infections between household members, but also the higher rates of clinic attendance by HIV-positive people, who are at increased risk of progression to disease following infection.

We parameterised our model to data from a high HIV prevalence, high TB, rural/peri-urban setting in KwaZulu-Natal, South Africa. The proportion of tuberculosis that results from transmission in clinics, and the impact of IPC interventions on community-wide TB incidence, is likely to vary by setting, depending on a range of factors. These include: the proportion of people’s contact time that occurs in clinics; the prevalence of HIV and other TB risk factors, and how clinic visiting and other contact behaviour varies between people with different risk factor profiles; and the number of clinic visits people with TB need to make before receiving a diagnosis. Social contact data from sub-Saharan Africa are limited^14^, however our estimate of the proportion of social contact that occurs in clinics falls with the range found by other studies^15 16^.

The social contact data used in our model were collected in 2019, before the start of the COVID-19 pandemic. Comparable social contact data were collected from the same study community in June-August 2020, during the pandemic, and suggested that reductions in contact time in clinics may have been smaller than reductions in other congregate locations^7^, possibly increasing the importance of transmission in clinics to overall disease burden during this period. The IPC interventions we simulated were also designed and parameterised before the start of the pandemic, and changing views on IPC may have changed the relative impact of the different interventions over longer time periods. For instance, the acceptability to patients of mask wearing may increase, increasing the coverage that can be achieved.

The main sources of uncertainty in our estimates come from three key inputs into the model: the proportion of contact time that occurs in clinics, the prevalence of TB in clinic attendees relative to the general population, and ventilation levels in clinics relative to other congregate settings.

Additional data collection in those three areas would be valuable, both in allowing us to reduce the uncertainty in our estimates, and in allowing similar estimates to be made for other settings.

There are a number of limitations to our work. Firstly, we do not explicitly consider infection to or from clinic staff. This may have led to us underestimating the proportion of disease that results from transmission in clinics, due to amplification of transmission in clinics by clinic staff, and to us slightly underestimated the impact of the interventions on community-wide TB incidence. The underestimates are likely to have been small however, as we estimate that only 7.1% (95% plausible range 2.3-16.7%) of all disease that results from transmission in clinics is in clinic staff, and contact time between clinic attendees and staff in clinics is much lower than contact time between clinic attendees. We also do not simulate children, as social contact data from children were not available. This will have had little effect on our estimates for adults, as the risk of *Mtb* transmission from children is low^17^, but means that we cannot estimate the proportion of disease in children that comes from transmission in clinics.

We do not consider the effects of risk factors other than HIV, such as diabetes. People with diabetes and some other risk factors are both likely to visit clinics more frequently, and are at increased risk of progression to disease following infection. By not including these risk factors, we may have underestimated the proportion of disease that results from transmission in clinics.

Finally, the representation of MDR in the model is relatively simple. We implicitly assume that with high coverage of Xpert MTB/RIF^18^, drug resistance is diagnosed for the majority of people at the same clinic visit as their TB is diagnosed. We therefore assumed that people with infectious MDR-TB spend no more time in clinics than people with infectious DS-TB, and so the proportion of TB from transmission in clinics does not vary by drug resistance status. We were not able to explore the effects of this assumption, due to the very low incidence of MDR-TB, and the need to use an IBM to accurately capture patterns of social contact behaviour. Future work should investigate if and if so, how, the proportion of MDR-TB that results from transmission in clinics varies from the proportion of DS-TB.

To conclude, we estimate that in the setting studied, 7.7% (4.0-14.2%) of tuberculosis in adults is acquired through transmission in PHC clinics, and that IPC interventions in clinics could reduce the total number of incident TB cases in the community in 2021-2030 by 3.4-8.0%. Given the relative ease of implementing IPC measures in clinics, compared to many other proposed TB control measures, we suggest that IPC measures in clinics should be considered to be ‘low hanging fruit’, and should be implemented both for their benefits to staff, but also for their likely effects on wider TB incidence and mortality.

## Data Availability

The mathematical model used in this work is available from https://github.com/NickyMcC/ClinicTransmissionModel The social contact data used in this work will be made available from https://datacompass.lshtm.ac.uk/ on publication of the manuscript.

https://datacompass.lshtm.ac.uk/

## Funding/acknowledgements

The support of the Economic and Social Research Council (IK) is gratefully acknowledged. The project is partly funded by the Antimicrobial Resistance Cross Council Initiative supported by the seven research councils in partnership with other funders including support from the GCRF. Grant reference: ES/P008011/1. NM is additionally funded the Wellcome Trust (218261/Z/19/Z). RGW is funded by the Wellcome Trust (218261/Z/19/Z), NIH (1R01AI147321-01), EDTCP (RIA208D-2505B), UK MRC (CCF17-7779 via SET Bloomsbury), ESRC (ES/P008011/1), BMGF (OPP1084276, OPP1135288 & INV-001754), and the WHO (2020/985800-0). RMGJH is funded by ERC (action number 757699). TAY receives an NIHR Academic Clinical Fellowship.

We would like to thank the social contact survey respondents, and all of the *Umoya omuhle* project team involved in the data collection. Full details are given in the supporting material.

## References

1. World Health Organization. Global tuberculosis report 2020, 2020.

2. World Health Organization. WHO guidelines on tuberculosis infection prevention and control: 2019 update: World Health Organization 2019.

3. Andrews JR, Morrow C, Walensky RP, et al. Integrating social contact and environmental data in evaluating tuberculosis transmission in a South African township. Journal of Infectious Diseases 2014;210(4):597–603.

4. McCreesh N, Grant AD, Yates TA, et al. Tuberculosis from transmission in clinics in high HIV settings may be far higher than contact data suggest. The International Journal of Tuberculosis and Lung Disease 2020;24(4):403–08.

5. Diaconu K, Karat AS, Bozzani F, et al. Systems interventions for improving TB infection prevention and control in South African primary care facilities

6. Gareta D, Baisley K, Mngomezulu T, et al. Cohort profile update: Africa Centre Demographic Information System (ACDIS) and population-based HIV survey. International journal of epidemiology 2021;50(1):33.

7. McCreesh N, Dlamini V, Edwards A, et al. Impact of social distancing regulations and epidemic risk perception on social contact and SARS-CoV-2 transmission potential in rural South Africa: analysis of repeated cross-sectional surveys. medRxiv 2020;2020.12.01.20241877 doi: https://doi.org/10.1101/2020.12.01.20241877

8. Newton SM, Brent AJ, Anderson S, et al. Paediatric tuberculosis. The Lancet infectious diseases 2008;8(8):498–510.

9. Govender I, Karat AS, Baisley K, et al. Prevalence of M. tuberculosis in sputum among clinic attendees compared with the surrounding community in rural South Africa: implications for finding the missing millions. 51st Union World Conference on Lung Health, 2020.

10. McCreesh N, Karat AS, Baisley K, et al. Modelling the effect of infection prevention and control measures on rate of Mycobacterium tuberculosis transmission to clinic attendees in primary health clinics in South Africa. medRxiv 2021;2021.07.26.21260835 doi: https://doi.org/10.1101/2021.07.26.21260835

11. Chartier Y, Pessoa-Silva C. Natural ventilation for infection control in health-care settings: World Health Organization 2009.

12. Health Systems Trust. The CCMDD story, 2019.

13. McCreesh N, White RG. An explanation for the low proportion of tuberculosis that results from transmission between household and known social contacts. Scientific reports 2018;8(1):5382.

14. Van Hoang T, Coletti P, Melegaro A, et al. A systematic review of social contact surveys to inform transmission models of close contact infections. bioRxiv 2018:292235.

15. Wood R, Racow K, Bekker L-G, et al. Indoor social networks in a South African township: potential contribution of location to tuberculosis transmission. PLoS One 2012;7(6):e39246.

16. McCreesh N, Looker C, Dodd PJ, et al. Comparison of indoor contact time data in Zambia and Western Cape, South Africa suggests targeting of interventions to reduce Mycobacterium tuberculosis transmission should be informed by local data. BMC Infectious Diseases 2016;16(1)

17. Middelkoop K, Bekker L-G, Morrow C, et al. Decreasing household contribution to TB transmission with age: a retrospective geographic analysis of young people in a South African township. BMC Infectious Diseases 2014;14(1):221. doi: 10.1186/1471-2334-14-221

18. National Health Laboratory Service. National Health Laboratory Service Annual Report 2018/19, 2019

