## Supporting material for "Estimating the contribution of transmission in primary healthcare clinics to community-wide TB disease incidence, and the impact of infection prevention and control interventions, in KwaZulu-Natal, South Africa"

### 1 Social contact data

#### 1.1 Methods

##### 1.1.1 Data collection

A social contact survey was conducted in the catchment areas of two primary health clinics in the southern section of the Africa Health Research Institute (AHRI) demographic surveillance area (DSA), between 28$^{th}$ March 2019 and 9$^{th}$ December 2019. 3090 adults (aged 18 and over) were sampled, stratified by local area.

Respondents were asked if they knew their HIV status. Respondents who reported being HIV-positive were asked if they were on anti-retroviral therapy (ART).

Respondent household size was extracted from existing DSA data.

Respondents were asked to list all indoor locations visited and transport used on an assigned day in the week before the survey. For each location visited (including their own home), they were asked for further details, including:

- What type of location it was (options included ‘own home’ and ‘clinic’)
- How long they spent there
- How many people (adults and children) were there, halfway through the time they were there
- How many of those people were children aged <15 years

For each use of transport reported, they were asked for further details, including:

- What type of transport it was
- How long the journey took
- How many people (adults and children) were on the vehicle at the start of the trip
- How many of those people were children aged <15 years

Respondents were also asked for additional details on their clinic visiting behaviour during the six months prior to the interview, including:

- The number of days on which they had visited a clinic for their own health in the past six months
- The number of days on which they had visited a clinic for on the behalf of someone else (e.g. to collect a prescription) in the past six months, not included any visits that were also made for their own health

- The number of days on which they had accompanied someone else to a clinic in the past six months, not including any visits that were also made for their own health and/or on behalf of someone else

Finally, respondents were asked when their last visit to a clinic was, and, if it was within the past two years, they were asked for the following information about their last visit:

- How long they spent at the clinic
- How many people (adults and children) were there, halfway through the time they were there
- How many of those people were children aged < 15

Further details of the social contact survey are given in McCreesh *et al*[1].

##### 1.1.2 Analysis

For each location visited on the assigned day, adult contact times were calculated as follows. Firstly, the number of adults present was calculated as the reported total number of people present, minus the reported number of children present. If this gave a value less than zero, it was set to missing. The number of adults present was then capped at 100, as above this value, it is unlikely that the respondent had sufficient contact with each adult present to allow transmission. The capped number of adults present was then multiplied by the duration of time that the respondent reported spending in the location, to give the adult contact time.

Estimates generated using the data on the respondent's last clinic visit were weighted by the reported number of clinic visits in the past six months.

Respondents who reported being HIV-positive were considered to be HIV-positive. Otherwise, respondents were considered to be HIV-negative/unknown.

#### 1.2 Results

##### 1.2.1 Recruitment

Of the 3090 people sampled for UO, 1723 (56%) were successfully contacted, 298 (10%) were dead or reported to have out-migrated, 1071 (35%) could not be contacted. Of those successfully contacted, 1704 (99%) completed an interview (Table S1).

| | | Sampled (%) | Contacted (%) | Dead or missing (%) | Responded (%) |
|---|---|---|---|---|---|
| **Sex** | Male | 1582 (51%) | 768 (45%) | 175 (59%) | 751 (44%) |
| | Female | 1508 (49%) | 955 (55%) | 123 (41%) | 953 (56%) |
| **Age group** | 18-29 | 1163 (38%) | 615 (36%) | 132 (44%) | 613 (36%) |
| | 30-49 | 1117 (36%) | 546 (32%) | 105 (35%) | 535 (31%) |
| | 50+ | 810 (26%) | 562 (33%) | 61 (20%) | 556 (33%) |
| **HIV status** | HIV negative or unknown | | | | 1210 (71%) |
| | HIV positive, not on ART | | | | 13 (1%) |
| | HIV positive, on ART | | | | 481 (28%) |
| **Household size** | 1-3 | | | | 293 (17%) |
| | 4-6 | | | | 426 (25%) |
| | 7-9 | | | | 429 (25%) |
| | 10+ | | | | 556 (33%) |
| **Total** | | 3090 | 1723 | 298 | 1704 |

**Table S1. Social contacts survey respondent characteristics**

##### 1.2.2 Time spent in own home

Respondents reported spending a mean of 18.8 (95% CI 18.5-19.1) hours per day in their own home. This varied little by sex, age group, HIV status, or household size (Table S2).

| | | Mean hours spent in own home per day (95% CI) |
|---|---|---|
| **Sex** | Male | 18.2 (17.8-18.7) |
| | Female | 19.2 (18.9-19.6) |
| **Age** | 18-29 | 18.1 (17.6-18.5) |
| | 30-49 | 18.3 (17.8-18.9) |
| | 50+ | 20.1 (19.6-20.5) |
| **HIV status** | Positive | 18.7 (18.4-19.0) |
| | Negative/Unknown | 19.1 (18.5-19.6) |
| **Household size** | 1-3 | 18.4 (17.7-19.1) |
| | 4-6 | 18.9 (18.3-19.4) |
| | 7-9 | 19.0 (18.4-19.5) |
| | 10+ | 18.8 (18.3-19.3) |
| **Overall** | | 18.8 (18.5-19.1) |

**Table S2. Mean reported time spent in own home, by sex, age, HIV status, and household size**

##### 1.2.3 Clinic visiting and contact time

###### *1.2.3.1 Frequency of clinic visiting*

Table S3 shows the estimated mean annual number of visits made to clinics, by sex, age, and HIV status, estimated from data on reported clinic visits in the past day, and in the past six months. Overall, there is little difference between the estimates calculated using the data collected using the two different recall durations. The exception to this is the estimates by sex, where there is a large difference in mean annual clinic visits by sex using the six-month recall data, but not the one-day recall data. However, the confidence intervals for the one-day recall estimates contain the estimated values for the six-month recall.

As there is no evidence that recall bias has had a large effect on the estimates, the six-month recall data are used to parameterise clinic visiting rates in the model, due to their greater precision.

| | | Mean annual clinic visits (95% CI) | |
|---|---|---|---|
| | | One-day recall | Six-month recall |
| **Sex** | Male | 7.8 (4.0-11.6) | 5.1 (4.7-5.4) |
| | Female | 7.7 (4.3-11.0) | 9.3 (8.8-9.7) |
| **Age** | 18-29 | 8.3 (4.0-12.7) | 6.7 (6.1-7.3) |
| | 30-49 | 8.9 (4.1-13.7) | 7.9 (7.4-8.5) |
| | 50+ | 5.9 (2.1-9.8) | 7.7 (7.2-8.2) |
| **HIV status** | Negative/Unknown | 6.7 (3.9-9.4) | 6.0 (5.7-6.4) |
| | Positive | 10.4 (5.0-15.7) | 10.8 (10.2-11.4) |
| **Overall** | | 7.7 (5.2-10.2) | 7.4 (7.1-7.7) |

**Table S3. Mean numbers of reported annual clinic visits by sex, age, and HIV status**

##### *1.2.3.2 Contact time*

Table S4 shows the mean adult contact hours per clinic visit, by sex, age, and HIV status, estimated from data on reported clinic visits in the past day, and in the past six months. Overall, there is little difference between the estimates calculated using the data collected using the two different recall durations. It is plausible, however, that the accuracy of recall for time spent in the clinic and numbers of people present falls fairly rapidly over time, and therefore the one-day recall estimates are used for estimating adult contact hours for input into the model.

| | | Mean adult contact hours per visit (95% CI) | |
|---|---|---|---|
| | | One-day recall | Last clinic visit* |
| **Sex** | Male | 150 (71-230) | 134 (121-147) |
| | Female | 131 (65-197) | 178 (165-190) |
| **Age** | 18-29 | 116 (47-185) | 163 (145-182) |
| | 30-49 | 179 (84-275) | 167 (149-185) |
| | 50+ | 112 (15-209) | 162 (149-175) |
| **HIV status** | Negative/Unknown | 138 (76-201) | 151 (138-164) |
| | Positive | 143 (55-232) | 182 (167-197) |
| **Overall** | | 140 (89-191) | 164 (155-174) |

**Table S4. Mean reported adult contact hours per clinic visit, by sex, age, and HIV status.** *Weighted by number of clinic visits in the past six months

##### 1.2.4 Contact in other locations

Other locations are defined as indoor locations other than clinics and the respondents' own homes, and transport.

Table S5 shows the mean adult contact hours in other locations, by sex, age, and HIV status.

| | | Mean contact hours per day (95% CI) |
|---|---|---|
| **Sex** | Male | 60 (51-70) |
| | Female | 58 (48-68) |
| **Age** | 18-29 | 75 (63-88) |
| | 30-49 | 49 (38-61) |
| | 50+ | 50 (37-62) |
| **HIV status** | Negative/Unknown | 64 (56-72) |
| | Positive | 47 (34-59) |
| **Overall** | | 59 (52-66) |

**Table S5. Mean reported contact hours per day in 'other' locations by sex, age, and HIV status**

### 2 Model description

#### 2.1 Key

Model parameter names are written in italics, with colour indicating whether the parameter is an input parameter, a parameter with a global model-wide value, calculated from input parameter(s) or other values, or an individual-level parameter, which can take a different value for each simulated person or household.

#### 2.2 Agents

Two types of agents were simulated in the model, people and households.

##### 2.2.1 People

The main state variables assigned to people in the model were:

- Unique ID – *person_ID*
- Age group – *age_group* (15-29, 30-49, 50-79)
- Sex – *sex* (male, female)
- Clinic visiting group – *clinic_group* (high, low)

- TB status – *TB_status* (uninfected, latent, smear+ disease, smear- disease, on treatment)
- TB strain – *TB_strain* (uninfected, drug sensitive (DS-TB), multidrug resistant (MDR-TB)
- Individual-level TB infectiousness – *infectiousness* (numeric, see section 'Individual-level variation in infectiousness)
- Location where last *Mtb* infection occurred – *infect_location* (uninfected, infected before creation, household, clinic, other location)
- HIV status – *HIV_status* (HIV-, HIV+ART-, HIV+ART+)

Other state variables were used to track individuals' histories in the model, for the purpose of creating model output.

##### 2.2.2 Households

Households were simulated as agents, for the purpose of grouping people into households with the desired size distributions. Households had the following state variables:

- Unique ID – *hh_ID*
- Desired household size – *desired_hh_size*
- Current household occupancy – *current_hh_size*

Other temporary household-level state variables were used to store information on the disease states of household members when estimating transmission probabilities in the household (see section '*Mtb* transmission – Household members')

###### *2.2.2.1 Household sizes*

Empirical data were available from the study population on the number of people aged 15+ years in each household. An exponential distribution was fitted to data on the cumulative proportion of households above each size, and the distribution was sampled from and rounded up to the nearest whole number to create desired household sizes in the model (Figure S1). Mean household sizes were similar between the model and the empirical data both from the perspective of households (model=3.64, data=3.97), and from the perspective of individuals (model=6.75, data=6.55).

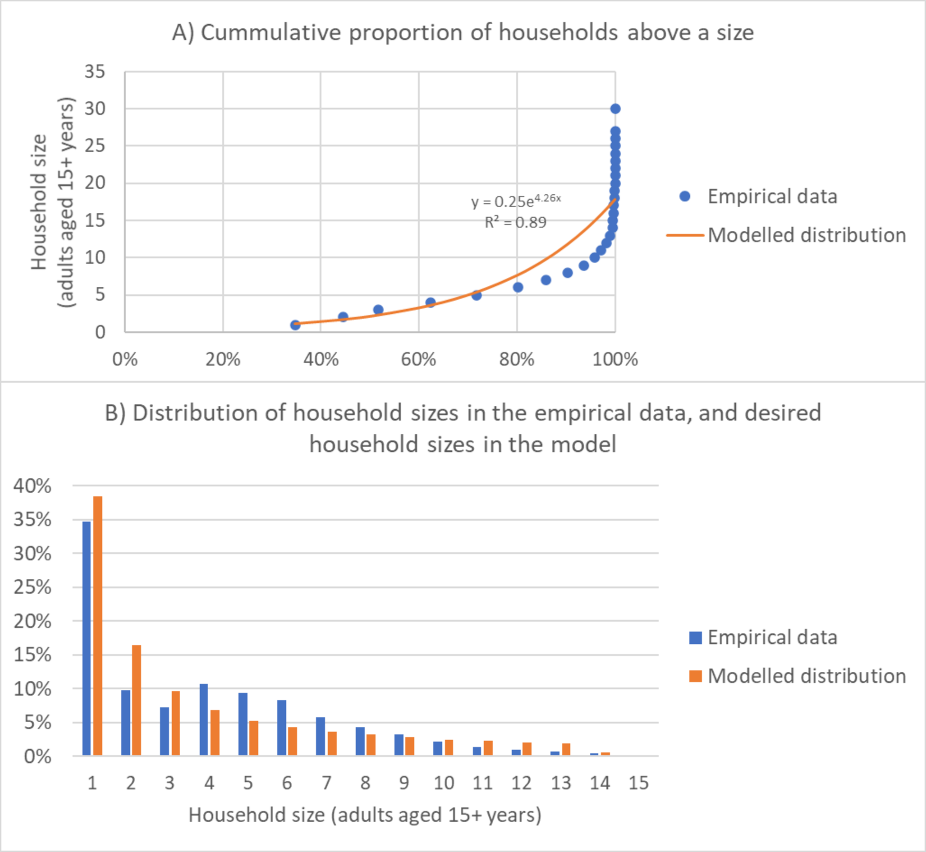

**Figure S1. Distribution of household sizes in the empirical data, and desired household sizes in the model.**

##### *2.2.2.2 Household occupancy tracking and formation*

To initialise the model, N empty households were created, where N = round (10,000 /*mean_hh_size*). Each empty household sampled a desired household size, *desired_hh_size*, from the exponential distribution (Figure S1), rounding up to the nearest whole number, and then created that number of people to populate the household, setting *current_hh_size* = *desired_hh_size*.

When people died, the household they were a member of reduced the value of *current_hh_size* by one. The household also added its *hh_ID* to the end of a list tracking households that are not at full occupancy.

When new people were created in the model, they checked the length of the list. If it was greater than one, the person joined the first household on the list. The household removed its *hh_ID* from the start of the list, and increased the value of *current_hh_size* by one.

If the length of the list was zero (i.e. there were no households that were not at full occupancy), then a new household was created, and the person joined it. The new household sampled a *desired_hh_size*, and if *desired_hh_size* > *1*, it added itself to the tracking list (*desired_hh_size* – 1) times.

#### 2.3 Model initialisation

To initialise the model, N empty households were created, where N = round (10,000 /*mean_hh_size*). Each empty household sampled a household size from an exponential distribution, rounding up to the nearest whole number (see section 'Household sizes' for details), and then created that number of people to populate the household. This gave an initial population size of approximately 10,000.

The newly created people were each assigned a *sex*, with a probability of 0.5 of being male and 0.5 of being female, and a *clinic_group* with and a probability of 0.5 of being 'high' and 0.5 of being 'low'. They were then assigned an *age_group*, with probabilities assigned by input parameters, and varying by sex; and an age, drawn from a uniform distribution between the minimum and maximum ages in their *age_group*. A random *infection_seed_proportion* were seeded with latent infection, with no risk of progression without reinfection. A random *tb_seed_proportion* were then seeded with TB disease, with *probability prop_smearpos_HIV0* becoming smear+ and the rest smear-.

The model was run with a constant population size for 100 years, and then a further 100 years with a growing population size, to allow the population age distribution and TB incidence and mortality to reach equilibrium. At that point, the model was considered to represent the year 2000, and realistic trends in HIV and TB were simulated from that point onwards.

#### 2.4 Model scheduling

The majority of events in the model were simulated using continuous time.

The two exceptions to this were the creation of new people, and the *Mtb* transmission process, which used a monthly time step.

#### 2.5 Model runs and calibration

The model was fitted by hand, by varying model input parameters until the model gave an acceptable fit to the fitting targets.

The model was run 2000 times for each fitted scenario and intervention, with the results averaged over the 2000 runs. Model outputs were outputted annually, giving mid-year values for cross-sectional count outputs, and end of year values for cumulative count outputs.

#### 2.6 Demography

Individuals were introduced into the model at age 15. People aged <15 were not modelled, as the risk of *Mtb* transmission from children is low[2], and contact data were not available from children from the study population.

During the initial run-in period, a constant population size of 10,000 was simulated. Each month, the number of people alive in the model was counted, and additional people created to restore the model population size to 10,000. After the initial run-in period, a constant birth rate per person alive was simulated, with the number of new people to be created each month equal to binomial(population size, *birth_rate*).

Exact age was tracked for each simulated individual; however, individuals were grouped into three age groups, 15-29 years, 30-49 years, and 50-79 years. A number of parameter values in the model varied by age group and sex: background mortality rates, HIV seeding proportions, HIV infection rates, and contact rates in clinics and 'other' locations.

There were four types of mortality in the model

- HIV mortality
- TB mortality
- Background mortality
- All individuals die upon reaching the age of 80 years

The background morality rates varied by age and sex, and were constant over time within each age group. TB and HIV mortality are described in the sections on TB and HIV.

##### 2.6.1 Fitting targets

The model was fitted to provincial-level data from KwaZulu-Natal on the estimated growth in population size between 2015 and 2019, the proportion of the population who are male in 2018, and the proportion of men and women in each of the three simulated age groups, by varying the simulated birth rate and age and sex specific background mortality rates. As in- and out-migration were not explicitly simulated, the background mortality rates were not designed to accurately reflect true (non-HIV and non-TB) mortality rates by age, but instead to also incorporate the effects of in- and out-migration on the population age distribution.

#### 2.7 Social contact

Three types of social contact were simulated in the model: contact between household members, contact occurring in clinics, and contact occurring in all other locations.

##### 2.7.1 Household members

In the model, it was assumed that each individual has *contact_time_each_hh_mem* = 572 hours of indoor contact with each member of their household each month (18.8 hours per day * 365.25 days / 12 months).

##### 2.7.2 Clinics

In line with the empirical data, the rate of clinic visiting in the model varied by sex and HIV/ART status, but not by age group. For each sex and HIV/ART status strata, 50% of the simulated population was assumed to be in a high clinic visiting group, and 50% in a low clinic visiting group. Clinic visiting rates in each group, for each strata, were determined by fitting a Poisson distribution to the data on the proportion of people in each strata who visited a clinic 0, 1 ,2-5 or 6+ times in the past six months, and the overall rate of clinic visiting in the strata, using a sum of least squares approach. Individuals changed between the high and low clinic visiting groups every six months with probability *clinic_rate_switch_prob*.

The rate of clinic visiting also varied for individuals with untreated TB disease (in the states smear-positive disease (smear+) and smear-negative disease (smear-)). Compared to individuals of the same sex, HIV/ART strata, and clinic visiting group, the rate of clinic visiting in people with untreated TB disease was increased by a factor of *increased_contact_time_clinics_tb*.

It was assumed in the model that all individuals had 140 adult contact hours on each clinic visit.

Individual clinic visits were not explicitly simulated in the model, instead each individual had a set amount of contact time in clinics each month (e.g. *contact_time_clinic_m_HIV01_low*), equal to the assumed mean number of clinic visits in a month (by sex, HIV/ART status, and clinic visiting group) multiplied by the mean contact time per visit.

##### 2.7.3 Other locations

Mean contact time in other locations in the model varied by sex, age group, and HIV/ART status, with mean contact time by group (e.g. *contact_time_other_m_age0_HIV01*) estimated using a regression model containing sex, age group, and HIV/ART status as categorical variables.

##### 2.7.4 Fitting targets

*increased_contact_time_clinics_tb* was varied to fit the model to empirical data from the study community in 2019 on the ratio of estimated prevalence of TB in clinic attendees relative to the general population[3]. The ratio was calculated from the model output as the proportion of all contact time in clinics in the model that was by people with smear+ or smear- TB, divided by the prevalence of smear+ or smear- TB in the whole model population, at the end of June 2019.

#### 2.8 Ventilation

Empirical data on ventilation rates in people's home in rural KwaZulu-Natal suggest mean absolute ventilation rates range from 110-274$m^3h^{-1}$ with windows and doors closed, 457-476 $m^3h^{-1}$ with windows open only, and 988-1187 $m^3h^{-1}$ with windows and doors open[4]. Empirical data from clinic waiting areas show large amount of variation in ventilation rates between different spaces, but they suggest that clinic spaces are generally better ventilated on average than people's homes[5]. We assumed in the model that the rate of transmission from a person with TB disease to a person without is 2.8 times higher in homes than in clinics. As the model is calibrated to an estimate of the proportion of disease that results from transmission between household members, however, the assumption made about ventilation rates in homes vs other spaces has little effect on the results (see Section 2.9.8).

Limited data were available on ventilation rates from other types of location, and showed large amounts of variation[6]. Nevertheless, rates for most locations were more in line with the higher ventilation rates found in clinic waiting areas than the lower rates found in people's houses. For this reason, we assumed in the main scenario in the model that the rate of transmission between a person with TB disease and a person without is the same in other locations as in clinics.

The effects of the assumptions made about ventilation rates in clinics and other locations were explored in a sensitivity analysis (See section 2.14 Uncertainty analysis).

#### 2.9 Tuberculosis

##### 2.9.1 Disease states

Each individual in the model was in one of five main TB states (uninfected, latent, smear+ disease, smear- disease, on treatment), with the latent infection state subdivided by time since infection (Figure S2).

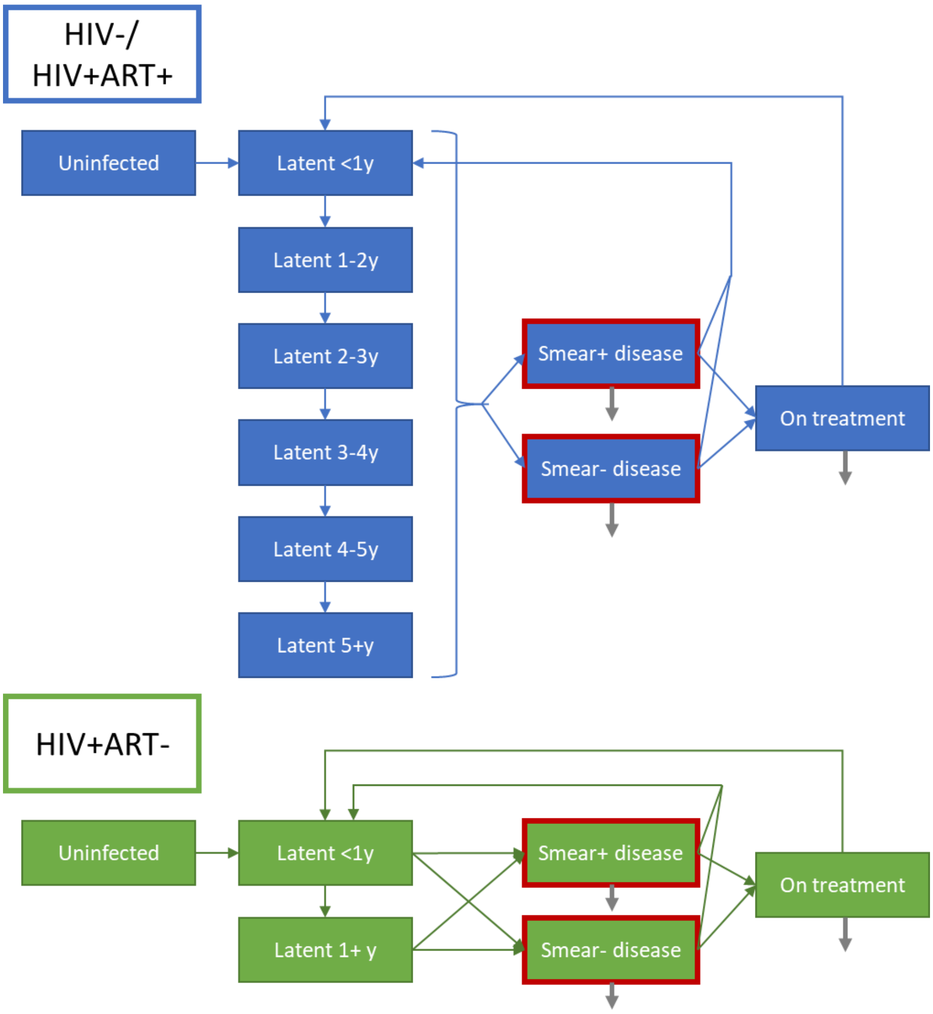

**Figure S2. Simulated TB states.** Blue and green boxes show the natural history for HIV-/HIV+ART+ and HIV+ART- individuals respectively. Grey arrows indicate tuberculosis mortality. Red outlines indicate infectious states.

##### 2.9.2 Drug resistance

Tuberculosis was simulated as drug sensitive (DS-TB) or multidrug resistant (MDR-TB). MDR-TB was seeded into the model in 2010 (*introduce_mdr_year*) by making simulated people in the model with *Mtb* infections (latent or active) set their resistance type to MDR-TB with probability *tb_seed_proportion_mdr*. MDR-TB was not introduced into the model earlier to prevent extinction of the strain when the model population size was lower.

Resistance type in the model effected the TB treatment duration. The treatment duration for DS-TB was always six months. For MDR-TB, it was 24 months for all people starting TB treatment before 2016, then 24 months with probability 0.3, and 11 months with probability 0.7[7 8].

TB treatment drop-out rates in the model also varied by resistance type (see Treatment).

##### 2.9.3 Disease progression

The rate of developing tuberculosis disease following infection depended on an individual's time since infection with *Mtb* and their HIV/ART status. For HIV- and HIV+ART+ people, the rate was highest in the first year, falling each year over the subsequent five years, and then lowest from five years following infection. For HIV+ART- people, the rate was highest in the first year following infection, and lower in all subsequent years.

The rate of developing disease also depended on the model year, being reduced by a factor of *decreased_tb_rates_late* for all simulated people in *change_TB_parameters_year* (see section changes in TB parameters over time), and for HIV+ART- people in *change_HIV1_parameters_year* (see section changes in HIV parameters over time).

Upon developing disease, HIV-, HIV+ART-, and HIV+ART+ people developed smear+ disease with *probability prop_smearpos_HIV0*, *prop_smearpos_HIV1*, and *prop_smearpos_HIV2* respectively. All other individuals developed smear- disease.

HIV-, HIV+ART-, and HIV+ART+ people with TB disease self-cured at rate *self_cure_rate_HIV0*, *self_cure_rate_HIV1*, and *self_cure_rate_HIV2* respectively. Upon self-cure, individuals re-entered the latent stage, resetting their time since infection back to zero.

##### 2.9.4 Treatment

Individuals with TB started treatment each month with probability *treatment_rate_HIV0* if HIV-, and *treatment_rate_HIV12* if HIV+. These rates took the value *treatment_rate_HIV0_early* and *treatment_rate_HIV12_early respectively* before *treatment_rate_change_year*, and *treatment_rate_HIV0_late* and *treatment_rate_HIV12_late* respectively afterwards.

After the year that ART was first introduced into the model, *ART_intro_year*, upon starting TB treatment, all HIV+ART- people became HIV+ART+.

Treatment lasted for *treatment_duration_DS* months if drug sensitive, and *treatment_duration_MDR* months if MDR. Individuals successfully finishing treatment re-entered the latent stage. Upon doing so, they reset their time since infection back to zero, reflecting the high rates of disease recurrence following treatment[9 10].

Individuals receiving TB treatment dropped out of treatment each month with probability *TB_treatment_dropout_rate_ DS* if they had DS-TB and, *TB_treatment_dropout_rate_MDR* if they had MDR-TB. Upon dropping out of treatment, they returned to active TB disease, with the same strain of disease (DS-TB or MDR-TB). Different TB treatment drop out rates by HIV status were not simulated, as empirical data showed little difference in treatment success by HIV status in South Africa[11].

##### 2.9.5 Mortality

TB mortality rates in the model depended on disease type (smear- or smear+), HIV/ART status, and whether someone was receiving treatment or not.

Among people not on treatment, the annual TB mortality rate was *TB_mortality_rate_smearpos_HIV0* (*TB_mortality_rate_smearneg_HIV0*) for HIV- with smear+ (smear-) disease, *TB_mortality_rate_smearpos_HIV1* (*TB_mortality_rate_smearneg_HIV1*) for HIV+ART- people with smear+ (smear-) disease, and *TB_mortality_rate_smearpos_HIV2* (*TB_mortality_rate_smearneg_HIV2*) for HIV+ART+ people with smear+ (smear-) disease.

When on treatment, the annual TB mortality rate was *TB_mortality_rate_treatment_DS* for people with DS-TB, and *TB_mortality_rate_treatment* for people with MDR-TB. Different TB mortality rates by HIV status while on TB treatment were not simulated, as empirical data showed little difference in treatment success by HIV status in South Africa[11].

##### 2.9.6 Prevalence of infection in 15-year olds

In 2013, 14.4% of 6-8 year olds were found to be infected with *Mtb* or to be on TB treatment in KwaZulu-Natal, giving an estimated annual rate of infection rate 2.1%[12]. Adjusting by reductions in estimated TB incidence between 2013 and 2018, and by increases in attack rates between childhood and adolescence[13], we estimated that around 24.2% of adolescents in KwaZulu-Natal in 2018 were infected with *Mtb*. Upon being created at the age of 15 years, people in the model therefore set their state to latent with probability 0.242. The remaining people were assumed to be uninfected.

In calculating rates of progression to active disease in individuals with *Mtb* infections at the point of their creation at age 15 in the model, we assigned them a time of infection, *time_of_infection*, from a uniform distribution covering the 15 years before their creation. Their rate of disease progression was then calculated using the same method as was used for people infected at ages >15 years. Progression to disease that occurred prior to the age of 15 was not included in the model.

*time_of_infection* was also used to determine, *prob_MDR_at_15*, the time-varying probability that individuals with existing infection at age 15 were infected with MDR *Mtb*. *prob_MDR_at_15* was set equal the proportion of the overall force of infection that was from individuals with MDR-TB at their assigned time of infection. For individuals created with a *time_of_infection* between *introduce_mdr_year – 15* and *introduce_mdr_year*, *prob_MDR_at_15* was set equal to *tb_seed_proportion_mdr*.

##### 2.9.7 Changes in TB natural history parameters over time

To reflect secular trends not captured by other time varying parameters in the model (for instance, improvements in nutrition and housing), a step change was modelled in *TB_parameter_change_year*. In *TB_parameter_change_year*, the simulated rate of *Mtb* transmission (*transmission_prob*), and the simulated rates of progression to TB disease following infection were reduced by a factor of *decreased_tb_rates_late*.

##### 2.9.8 Fitting targets

The model was fitted to a range of TB incidence, mortality, and treatment outcome estimates (see section 'Modelling fitting targets').

The model was also fitted to the central value of a range of estimates for the proportion of disease that results from transmission between household members in sub-Saharan African countries[14]. This was done by varying the degree of individual variation in infectiousness between people with tuberculosis, with higher levels of variation leading to a lower proportion of disease resulting from transmission between household members.

#### 2.10 *Mtb* transmission

*Mtb* transmission in the model was scheduled on a monthly time step. Three transmission 'locations' were simulated, with transmission in each location simulated in turn each month: transmission between household members, transmission in clinics, and transmission in other indoor locations (including transport). Random mixing was assumed in clinics and in other locations.

In all locations, the parameter *transmission_prob* determined the baseline probability of transmission per minute contact between each uninfected or latent person and each person with smear+ or smear- TB. *transmission_prob* took the value *transmission_prob_early* before *TB_parameter_change_year*, and *transmission_prob_early* * *decreased_tb_rates_late* afterwards.

The baseline *transmission_prob* was then adjusted for a number of factors:

- The simulated ventilation level in the location. The effect of ventilation levels on the rate of transmission is described in the section 'Ventilation'.
- The smear status of the person with TB. We assumed that people with smear- disease are 78% less infectious than people with smear+ TB[15].
- Whether the exposed person was uninfected or latent, and their HIV/ART status. We assumed that latent infection provides 72% protection against reinfection in HIV- people[16], with lower levels of protection in HIV+ART- people, and intermediate levels of protection in HIV+ART+ people.

- The individual-level infectiousness of the person with TB (household transmission only) (see section 'Individual-level variation in infectiousness).

##### 2.10.1 Individual-level variation in infectiousness

Individuals in the model had an individual level of infectiousness, *infectiousness*. This was sampled at birth for each simulated person from a gamma distribution with mean = 1 and variance = *infectiousness_var*.

The *infectiousness* parameter was assumed to incorporate the effects of all factors that have an effect on the infectiousness of a person with TB, with the exception of whether the disease is smear+ or smear-.

Individual-level variation in infectiousness was simulated when determining *Mtb* transmission between household members, because the variation acts to reduce the rate of transmission between highly regular contacts such as household members, through increasing the effects of saturation[14]. Not incorporating this variation would therefore have resulted in an unrealistically high proportion of disease in the model coming from transmission between household members.

Individual-level variation in infectiousness was not used in the model when determining *Mtb* transmission in clinics and other locations. Instead, the overall mean value of *infectiousness*, 1, was used for all people. This reduced model stochasticity, speeding up the model fitting process, and meaning that far fewer model runs needed to be done per final scenario and intervention. As random mixing was simulated in both clinics and other locations, this had no effect on the average proportion of disease that results from transmission in clinics and other locations in the model.

##### 2.10.2 Household members

To simulate transmission between household members, the number of people with smear+ DS-TB and MDR-TB, and smear- DS-TB and MDR-TB, in each household were counted ($N_{sr}$, where s=0 indicates smear- disease and s=1 indicates smear+ disease, and where r=0 indicates DS-TB and r=1 indicates MRD-TB), and the mean value of *infectiousness* in household members with smear+ DS-TB and MDR-TB and smear- DS-TB and MDR-TB was calculated for each household ($I_{sr}$). If no household members had the corresponding type of disease, then $I_{sr}$ was set to zero.

For each susceptible or latent individual in the household, the probability of infection each month was calculated as:

$$1 - \prod_{s=0}^{1}\prod_{r=0}^{1}(1 - transmission\_prob \times I_{sr} \times ventilation\_weight\_home \times reinfection\_relative\_risk \times W_s)^{N_{sr} * contact\_time\_each\_hh\_mem})$$

Where:

- *reinfection_relative_risk* = 1 if the individual was uninfected, *reinfection_relative_risk_HIV0* if they were HIV- and latently infected, *reinfection_relative_risk_HIV1* if they were HIV+ART- and latently infected, and *reinfection_relative_risk_HIV2* if they were HIV+ART+ and latently infected
- $W_s$ = 1 when s = 1, and $W_s$ = *reduced_transmission_smearneg* when s = 0

The probability that people infected with *Mtb* from transmission from a household member were infected with an MDR strain was calculated as:

$$(\sum_{s=0}^{1} N_{s1} \times I_{s1} \times W_s)/(\sum_{s=0}^{1}\sum_{r=0}^{1} N_{sr} \times I_{sr} \times W_s)$$

##### 2.10.3 Clinics

Each month, the total contact number of people in each class was counted, with class defined as the 60 strata generated by all combinations of:

- Sex (male, female)
- HIV/ART status (HIV-, HIV+ART-, HIV+ART+)
- Clinic visiting group (high, low)
- TB status (smear+ DS-TB, smear- DS-TB, smear+ MDR-TB, smear- MDR-TB, non-infectious (all other TB states))

The total contact time in clinics by people in each class was then calculated, by multiplying the number of people by the mean contact time per person. For people with smear+ and smear- TB, mean contact time was higher by a factor of *increased_contact_time_clinics_tb*, compared to other people in the same sex, HIV/ART, and clinic visiting strata.

Finally, the proportion of all contact time in clinics that were with someone with smear+ DS-TB, smear+ MRD-TB, smear- DS-TB, and smear- MRD-TB was calculated ($P_{sr}$, where s=0 indicates smear- disease and s=1 indicates smear+ disease, and where r=0 indicates DS-TB and r=1 indicates MRD-TB).

For each susceptible or latent individual in the model, the probability of infection each month from transmission in clinics was then calculated as:

$1 - \prod_{s=0}^{1}\prod_{r=0}^{1}(1 \; - transmission\_prob \times int\_RR\_trans\_clinics \times ventilation\_weight\_clinics \times reinfection\_relative\_risk \times W_s)^{P_{sr} \, * \, contact\_time\_clinics \times int\_RR\_contact\_clinics})$

Where:

- *reinfection_relative_risk* = 1 if the individual was uninfected, *reinfection_relative_risk_HIV0* if they wer HIV- and latently infected, *reinfection_relative_risk_HIV1* if they were HIV+ART- and latently infected, and *reinfection_relative_risk_HIV2* if they were HIV+ART+ and latently infected.
- $W_s$ = 1 when s = 1, and $W_s$ = *reduced_transmission_smearneg* when s = 0.
- *contact_time_clinics* was equal to the mean monthly contact time in clinics for someone of the individual's class.
- *int_RR_trans_clinics* = 1 until 2021 in all scenarios, and took different values from then in some intervention scenarios (see 'Interventions').
- *int_RR_contact_clinics* = *int_RR_contact_clinics_HIV01* if the individual was HIV- or HIV+ART-, and *int_RR_contact_clinics* = *int_RR_contact_clinics_HIV2* if the individual was HIV+ART+. *int_RR_contact_clinics_HIV01* = *int_RR_contact_clinics_HIV2* = 1 until 2021 in all scenarios, and took different values from then in some intervention scenarios (see 'Interventions').
- 1 until 2021 in all scenarios, and took different values from then in some intervention scenarios (see 'Interventions').

The probability that people infected with *Mtb* from transmission in clinics were infected with an MDR strain was calculated as:

$$(\sum_{s=0}^{1} P_{s1} \times W_s)/(\sum_{s=0}^{1}\sum_{r=0}^{1} P_{sr} \times W_s)$$

##### 2.10.4 Other locations

Each month, the total contact number of people in each class was counted, with class is defined as the 90 strata generated by all combinations of:

- Sex (male, female)
- HIV/ART status (HIV-, HIV+ART-, HIV+ART+)
- Age group (15-29, 30-49, 50-79)

- TB status (smear+ DS-TB, smear- DS-TB, smear+ MDR-TB, smear- MDR-TB, non-infectious (all other TB states))

The total contact time in other location by people in each class was then calculated, by multiplying the number of people by the mean contact time per person.

Finally, the proportion of all contact time in other locations that was with someone with smear+ DS-TB, smear+ MRD-TB, smear- DS-TB, and smear- MRD-TB was calculated ($P_{sr}$, where s=0 indicates smear- disease and s=1 indicates smear+ disease, and where r=0 indicates DS-TB and r=1 indicates MRD-TB).

For each susceptible or latent individual in the model, the probability of infection each month from transmission in other locations was then calculated as:

$$1 - \prod_{s=0}^{1}\prod_{r=0}^{1}(1 - transmission\_prob \times ventilation\_weight\_other \times reinfection\_relative\_risk \times W_s)^{P_{sr} * contact\_time\_other})$$

Where:

- *reinfection_relative_risk* = 1 if the individual was uninfected, *reinfection_relative_risk_HIV0* if they were HIV- and latently infected, *reinfection_relative_risk_HIV1* if they were HIV+ART- and latently infected, and *reinfection_relative_risk_HIV2* if they were HIV+ART+ and latently infected.
- $W_s$ = 1 when s = 1, and $W_s$ = *reduced_transmission_smearneg* when s = 0
- *contact_time_other* was equal to the mean monthly contact time in other locations for someone of the individual's class.

The probability that people infected with *Mtb* from transmission in other locations were infected with an MDR strain was calculated as:

$$(\sum_{s=0}^{1} P_{s1} \times W_s)/(\sum_{s=0}^{1}\sum_{r=0}^{1} P_{sr} \times W_s)$$

#### 2.11 HIV/ART

Three HIV states were simulated in the model: HIV-, HIV+ART-, and HIV+ART+.

HIV was introduced into the model in 2000, by seeding a set proportion of each age group and sex at random with HIV. People created in the model at age 15 years were all HIV-. From the introduction of HIV in the model in 2000, HIV- people became HIV+ART- at a rate that varied by age group and sex.

To capture changes in estimated and projected HIV prevalence over time, the value of the HIV incidence parameters for each age group and sex changed twice in the model, in *HIV_inc_change_year1* and *HIV_inc_change_year2*.

ART was introduced in the model in 2005. From the introduction of ART, HIV+ART- people became HIV+ART+ at a rate that varied by sex. To capture changes in estimated ART coverage over time, the values of the ART start rates in the model were changed in *ART_start_rate_change_year*.

From *ART_start_rate_change_year*, all HIV+ART- people starting TB treatment were made HIV+ART+.

HIV mortality was simulated as a constant rate of (non-TB) HIV-related mortality for all HIV+ART- people (*HIV1_mortality_rate*), and all HIV+ART+ people (*HIV2_mortality_rate*),

##### 2.11.1 Effects on TB

HIV and ART status effected a number of TB-related rates and probabilities in the model:

- TB mortality rates
- Rates of progression to disease
- Self-cure rates
- Protection against reinfection from being latently infected
- Probability of developing smear+ disease
- Contact rates

The effects of HIV and ART on parameter values are described in more details in the relevant sections, and the parameter ranges shown in the section ‘Input parameters’. When someone became HIV+ or started ART, the values of all of their rates change immediately.

##### 2.11.2 Changes in HIV parameters over time

CD4 counts for HIV+ART- people were not explicitly simulated, with HIV+ART- being simulated as a single, homogenous group, varying only with age group and sex. As ART coverage increased over time in South Africa, however, the average CD4 count of people not on ART is likely to have risen, and the impact on TB natural history of being HIV+ART- is likely to changed.

To allow the effects of increased ART coverage on TB to be adequately captured in the model, enabling the model to be fitted to trends in TB incidence over time, a step change in the values of certain HIV related parameters was simulated, starting in *change_HIV1_parameters_year*.

From *change_HIV1_parameters_year*, the degree of protection that latent infection gave against reinfection in HIV+ART- people, *reinfection_relative_risk_HIV1*, was increased from *reinfection_relative_risk_HIV1_early* to *reinfection_relative_risk_HIV1_late*, and the rate of developing disease in more than one year following infection in HIV+ART- people was decreased from *develop_tb_reactivation_rate_HIV1_early* to *develop_tb_reactivation_rate_HIV1_late*. From *TB_parameter_change_year*, these rates were also decreased by *decreased_tb_rates_late* (see section 'Changes in TB natural history parameters over time' and Table S6). As the rate of developing disease in the first year following infection in HIV+ART- people was calculated relative to the rate in subsequent years in the model, this also decreased the rate in the first year following infection.

| Order of two parameter value change years | First time period | | Second time period | | Third time period | |
|---|---|---|---|---|---|---|
| | Time range | Parameter value | Time range | Parameter value | Time range | Parameter value |
| *change_hiv1_parameters_year* < *TB_parameter_change_year* | Start to *change_hiv1_parameters_year* | *reinfection_relative_risk_HIV1_early* | *change_hiv1_parameters_year* to *TB_parameter_change_year* | *reinfection_relative_risk_HIV1_late* | *TB_parameter_change_year* to end | *reinfection_relative_risk_HIV1_late* * *decreased_tb_rates_late* |
| *change_hiv1_parameters_year* > *TB_parameter_change_year* | Start to *TB_parameter_change_year* | *reinfection_relative_risk_HIV1_early* | *TB_parameter_change_year* to *change_hiv1_parameters_year* | *reinfection_relative_risk_HIV1_early* * *decreased_tb_rates_late* | *change_hiv1_parameters_year* to end | *reinfection_relative_risk_HIV1_late* * *decreased_tb_rates_late* |
| *change_hiv1_parameters_year* = *TB_parameter_change_year* | Start to *change_hiv1_parameters_year*/ *TB_parameter_change_year* | *reinfection_relative_risk_HIV1_early* | *change_hiv1_parameters_year*/ *TB_parameter_change_year* to end | *reinfection_relative_risk_HIV1_late* * *decreased_tb_rates_late* | NA | NA |

**Table S6. Value taken by the model parameter *reinfection_relative_risk_HIV1*, over time, depending on the relative values of the parameter *change_hiv1_parameters_year* and *TB_parameter_change_year*.**

##### 2.11.3 Fitting targets

The model was fitted to a range of HIV prevalence and ART coverage targets, based on empirical estimates from the study population[17]. These are described in full in the section 'Fitting targets'.

In addition to this, the model was fitted to estimated future trends in HIV prevalence and ART coverage by sex, from provincial HIV model (Thembisa) estimates[18 19]. As the Thembisa estimates were for the province as a whole, and the model was fitted to historic trends from the study population, the model was fitted to estimates *changes* in HIV prevalence and ART coverage by sex between 2020 and 2030, rather than the absolute estimates.

#### 2.12 Interventions

Seven potential infection control interventions had been identified in qualitative research and system dynamics modelling exercises conducted as part of the *Umoya omuhle* project[20]. The effect of the interventions on patient contacts and infection risk in clinics were estimated in previous modelling work, using a within-clinics model that simulated the flow of patients through clinics, and ventilation rates and infection risk in clinic waiting areas[21]. The interventions were:

1) **Opening windows and doors.** Ensuring windows and doors in waiting areas are kept open at all times. This was implemented in the within-clinics model through increasing simulated ventilation rates
2) **Simple clinic retrofits.** Building retrofits are changes to the building to improve ventilation rates. This could include installing lattice brickwork or whirlybird fans. Due to the large amount of variation between clinic spaces in the types of building retrofits that would be suitable, and the lack of sufficient data on the effects of the retrofits on ventilation rates in different types of spaces, we did not model specific retrofits or packages of retrofits. Instead, in the within clinics model, we simulated an undefined package of retrofits that are sufficient to increase air changes per hour to a minimum of 12 in all rooms, chosen in line with WHO guidelines[22 23]
3) **Ultraviolet Germicidal Irradiation (UVGI) system.** We assumed in this intervention that appropriate and well maintained UVGI systems are installed in all indoor clinic waiting areas. This was implemented in the within-clinics model through an additional quanta clearance rate, equivalent to a ventilation rate of 24 ACH (95% CI 9.9-62)[24].
4) **Surgical mask wearing by patients.** We simulated a scenario where 70% of patients wear surgical masks 90% of the time. Masks were assumed in the within-clinics model to

reduce the rate of quanta production by 75% (95% CI 56-85%)[25], and have no effect on rate of infection for the person wearing the mask[26].

5) **Increasing CCMDD coverage.** South Africa's Central Chronic Medicine Dispensing and Distribution (CCMDD) programme is designed to allow patients with stable chronic health conditions to collect their medicines from convenient locations, such as local pharmacies[27]. This means that they do not need to queue at clinics unnecessarily. The purpose of this intervention was to increase the utilisation of CCMDD and similar programmes by eligible patients, and to ensure that pick-up points do not require patients to queue at clinics. We assumed that 92% (95% CI 84-95%) of patients could have their ART appointments reduced to once every 6 months[28], and that the remaining 8% of people need monthly ART appointments. This was implemented in the within-clinics model through removing 31% (IQR 22-34%) of ART patients, chosen at random each model run.
6) **Queue management system with outdoor waiting areas.** Empirical data show that clinic waiting areas are often crowded, and that in many clinics patients wait in unsuitable areas such as corridors[29]. This is partly due to patient concerns that if they wait in other areas, they may not hear their name being called, and may miss their turn. This intervention therefore combined a large, covered outdoor waiting area with a queue management system, such as numbered tickets or an electronic tracking system. We assumed in the within-clinics model that only a small number of patients were allowed to wait inside the clinic, with the rest waiting in a large, covered, outdoor waiting area, with a very high ventilation rate of 52-70 ACH[30].
7) **Appointment systems.** In this intervention, we simulated a date-time appointment system to reduce clinic overcrowding, through spacing out the arrival times of patients in the within-clinics model.

The estimated effects of the interventions on patient contacts and infection risk in clinics from the within-clinics model were used to parameterise the effects of the interventions in this model, allowing their wider effects on community-level disease incidence to be estimated. The interventions were implemented through changing parameter values, starting in 2021 (see Table S7).

The 'best estimates' of intervention effects in this model were informed by the median impacts from the within-clinics model. The minimum and maximum estimates were informed by the interquartile ranges from the within-clinics model. The interquartile range was used, rather than the full range, as the most extreme effects from the within-clinics model were assumed to reflect day to day variation, rather than genuine uncertainty in intervention effects.

| Intervention | Parameters changed | Parameter description | Simulated value (from 2021) | | |
|---|---|---|---|---|---|
| | | | **Minimum effect** | **Best estimate** | **Maximum effect** |
| **Windows and doors** | *int_RR_trans_clinics* | Modifier of risk of infection per minute contact occurring in clinics | 0.75 | 0.45 | 0.28 |
| **Retrofits** | *int_RR_trans_clinics* | | 0.84 | 0.55 | 0.36 |
| **UVGI** | *int_RR_trans_clinics* | | 0.36 | 0.23 | 0.15 |
| **Masks** | *int_RR_trans_clinics* | | 0.58 | 0.53 | 0.50 |
| **Queue management and outdoor waiting area** | *int_RR_trans_clinics* | | 0.24 | 0.17 | 0.12 |
| **CCMDD** | *int_RR_contact_clinics_HIV2*<br>*int_RR_contact_clinics_HIV01* | Modifier of mean contact hours in clinics for people who are HIV+ART+ and HIV- or HIV+ART- respectively | HIV2: 0.91<br>HIV01: 0.92 | HIV2: 0.72<br>HIV01: 0.87 | HIV2: 0.58<br>HIV01: 0.81 |
| **Appointment system** | *int_RR_contact_clinics_HIV2*<br>*int_RR_contact_clinics_HIV01* | Modifier of mean contact hours in clinics for people who are HIV+ART+ and HIV- or HIV+ART- respectively | Both: 0.55 | Both: 0.38 | Both: 0.25 |

**Table S7. Simulated intervention effects.** Parameters changed in each intervention, and the values simulated

#### 2.13 Results calculations

When calculating the proportion of disease that resulted from transmission in clinics in the model, simulated individuals who developed disease from an infection that occurred before the age of 15 years were not included, as their location of infection could not be determined.

Intervention effects on TB incidence and mortality were calculated as relative changes in rates, compared to a scenario where no interventions are simulated. As the simulated proportion of people created in the model at age 15 years who had a latent infection is constant over time, simulated individuals who developed or died from TB disease from an infection that occurred before the age of 15 years were not included when estimating intervention effects on TB incidence and mortality.

#### 2.14 Uncertainty analyses

A number of univariate sensitivity analyses were conducted:

- **Proportion of outside-household contact time occurring in clinics (clinic contact time).** From the social contact data, overall, we estimated that 5.3% (95% CI 2.8-8.0%) of contact time that occurs outside respondents' own homes occurs in clinics (weighted to model population size by sex and HIV/ART status in 2019). In the sensitivity analysis, we explored the effect of multiplying all of the clinic contact parameters by 0.53 (=2.8/5.3) and 1.51 (=8.0/5.3). The simulated clinic contact times are shown in Table S8.
- **Prevalence of TB in clinic attendees relative to the general population (TB in clinics).** In the main scenario, the model was fitted to a prevalence of TB in clinic attendees relative to the community prevalence of 1.86. In the sensitivity analysis, the model is fit to the upper bounds of the empirical 95% confidence interval (1.1-3.1)[3]. Fitting to the lower bound would have required the value of *increased_contact_time_clinics_tb* to be less than one. In other words, it would have required simulating a lower rate of clinic visiting in people with TB compared to people without, controlling for sex and HIV/ART status. This was considered to be implausible, therefore *increased_contact_time_clinics_tb* = 1 was used as the lower bound.
- **Proportion of disease from household transmission (Household transmission).** In the main scenario, we fitted the model to 13.5% of disease resulting from transmission between household members. In the sensitivity analysis, the model was fitted to 8% and 19%[14] of disease resulting from transmission between household members. This was achieved primarily by changing the value of *infectiousness_var*.

- **Ventilation rates in clinics (Clinic ventilation).** In the main scenario, mean ventilation rates were assumed to be the same in clinics as in other locations, with *ventilation_weight_clinic* = *ventilation_weight_other* = 1. In the sensitivity analysis, the value of *ventilation_weight_clinic* was changed to 0.5 and to 2.
- **Movement between high and low clinic visiting groups (Clinic risk groups).** As the social contact survey collected data on number of clinics visits over a six-month period only, we were unable to distinguish the extent that differences in clinic visiting rates between people of the same sex and HIV/ART status were due to long-term, stable differences vs shorter term fluctuations in clinic use. In the main scenario, we simulated people switching between clinic visiting risk groups every six months with probability *clinic_rate_switch_prob* = 0.25. In the sensitivity analysis, we simulated people switching with probability 0 and 0.5.
- **Clinic visiting rates by HIV+ART- people, relative to HIV- people (HIV+ART- clinic visiting).** In the social contact data collection, only 13 people reported being HIV+ART-. In addition, HIV-status was self-reported, and we could therefore not accurately distinguish between HIV- and undiagnosed HIV+ people, particularly when the reported date of the last HIV-test was not recent. We therefore had no empirical data on rates of clinic visiting in HIV+ART- people. In the main scenario, we assumed that the rates are the same in HIV+ART- and in HIV- people, and determined the rates from the empirical data for all people who did not report being on ART. In the sensitivity analysis, we assumed that rates in HIV+ART- people are half that of HIV- people, and that rates in HIV+ART- people are the same as for HIV+ART+ people. In both scenarios, we also adjusted the HIV- clinic visiting rates to keep the overall mean clinic visiting rates in 2020 for HIV+ART- and HIV- people constant. The simulated clinic contact times are shown in Table S8.
- **Future HIV incidence.** Estimated future trends in HIV incidence were taken from the projections from a provincial-level HIV model, Thembisa[18 19], with the model fitted to the estimated change in HIV prevalence in men and women between 2020 and 2030. While Thembisa did provide 95% limits for its estimates, we considered them to be unrealistically narrow. For instance, the 95% limits for the projected prevalence of HIV in men aged 15-49 in 2030 was 11.4-12.3%. In the sensitivity analysis, we therefore chose to simulate relative changes in HIV incidence by sex from 2020 compared to the preceding time period, that were 50% lower and 150% higher than the simulated changes in the main scenario.

In all sensitivity analyses, the model was recalibrated to the same fitting targets (with the exception of the targets explicitly changed in the sensitivity analysis).

| | Best | Clinic contact time | | HIV+ART- clinic visiting | |
|---|---|---|---|---|---|
| | | Low | High | Low | High |
| *contact_time_clinic_m_HIV0_low* | 493 | 256 | 737 | 520 | 143 |
| *contact_time_clinic_m_HIV1_low* | 493 | 256 | 737 | 260 | 3468 |
| *contact_time_clinic_m_HIV2_low* | 3468 | 1803 | 5185 | 3468 | 3468 |
| *contact_time_clinic_f_HIV0_low* | 2322 | 1207 | 3472 | 2483 | 1435 |
| *contact_time_clinic_f_HIV1_low* | 2322 | 1207 | 3472 | 1242 | 8276 |
| *contact_time_clinic_f_HIV2_low* | 8276 | 4302 | 12374 | 8276 | 8276 |
| *contact_time_clinic_m_HIV0_high* | 5507 | 2863 | 8234 | 5812 | 5167 |
| *contact_time_clinic_m_HIV1_high* | 5507 | 2863 | 8234 | 2906 | 8400 |
| *contact_time_clinic_m_HIV2_high* | 8400 | 4367 | 12560 | 8400 | 8400 |
| *contact_time_clinic_f_HIV0_high* | 8609 | 4475 | 12872 | 9206 | 8658 |
| *contact_time_clinic_f_HIV1_high* | 8609 | 4475 | 12872 | 4603 | 8276 |
| *contact_time_clinic_f_HIV2_high* | 8276 | 4302 | 12374 | 8276 | 8276 |

**Table S8. Simulated clinic contact time per month in the best scenario, and clinic contact time and HIV+ART- clinic visiting scenarios.** Values in all other uncertainty analysis scenarios are the same as in the best scenario.

#### 2.15 Input parameters

| Name | Description | Value/range | Source |
|---|---|---|---|
| | **Tuberculosis parameters** | | |
| *tb_seed_proportion* | Proportion of people seeded with TB at the start of the model run | 0.005 | NA. Model allowed to reach equilibrium before output produced |
| *infection_seed_proportion* | Proportion of people seeded with latent Mtb infection at the start of the model run | 0.7 | NA. Model allowed to reach equilibrium before output produced |
| *transmission_prob_early* | Baseline rate of *Mtb* transmission per minute meeting time (before adjustment) | 0-1 | Varied to fit data |
| *TB_parameter_change_year* | Year from which the value of *transmission_prob* and simulated disease progression rates are changed | 2007-2018 | From year in which estimated TB incidence starts to decline, to final TB incidence fitting year |
| *decreased_tb_rates_late* | Multiplier for transmission rate and disease progression rates from *TB_parameter_change_year* | 0-1 | Varied to fit data |
| *reduced_transmission_smearneg* | Lower transmission rate with smear- disease, relative to smear+ | 0.22 | Houben[15] |

| | | | |
|---|---|---|---|
| *reinfection_relative_risk_HIV0* | Reduced probability of transmission to people with latent infections, relative to uninfected people (HIV-) | 0.28 | Dowdy and Chaisson[16] |
| *reinfection_relative_risk_HIV1_early* | Reduced probability of transmission to people with latent infections, relative to uninfected people, prior to *change_HIV1_parameters_year* (HIV+ART-) | > 0.75 | Dowdy and Chaisson[16] |
| *reinfection_relative_risk_HIV1_late* | Reduced probability of transmission to people with latent infections, relative to uninfected people, from *change_HIV1_parameters_year* (HIV+ART-) | > *reinfection_relative_risk_HIV2*<br>< 0.75 | Dowdy and Chaisson[16] |
| *reinfection_relative_risk_HIV2* | Reduced probability of transmission to people with latent infections, relative to uninfected people (HIV+ART+) | > *reinfection_relative_risk_HIV0*<br>< *reinfection_relative_risk_HIV1_late* | |
| *infectiousness_var* | The between-individual variance in infectiousness | >0 | Varied freely to fit data |
| *self_cure_rate_HIV0* | The annual rate of self-cure for HIV- people | 0.2 | Estimated from Menzies *et al*[31] |
| *self_cure_rate_HIV1* | The annual rate of self-cure for HIV+ART- | 0.08 | Estimated from Menzies *et al*[31] |
| *self_cure_rate_HIV2* | The annual rate of self-cure for HIV+ART+ | 0.14 | Estimated from Menzies *et al*[31] |
| *TB_mortality_rate_smearpos_HIV0* | Annual rate of mortality from smear+ pulmonary or extrapulmonary TB for HIV- people | 0.335-0.449 | Ragonnet *et al* (2020)[32] |

| | | | |
|---|---|---|---|
| *TB_mortality_rate_smearpos_HIV1* | Annual rate of mortality from smear+ pulmonary or extrapulmonary TB for HIV+ART- people | > *TB_mortality_rate_smearpos_HIV0* | |
| *TB_mortality_rate_smearpos_HIV2* | Annual rate of mortality from smear+ pulmonary or extrapulmonary TB for HIV+ART+ people | Between 0.16 and 0.91 times *TB_mortality_rate_smearpos_HIV1*, and ≥ *TB_mortality_rate_smearpos_HIV0* | Dheda *et* al (2004)[33] and Lawn *et al* (2009)[34] |
| *TB_mortality_rate_smearneg_HIV0* | Annual rate of mortality from smear- pulmonary TB for HIV- people | 0.017-0.035 | Ragonnet *et al* (2020)[32] |
| *TB_mortality_rate_smearneg_HIV1* | Annual rate of mortality from smear- pulmonary TB for HIV+ART- | > *TB_mortality_rate_smearneg_HIV0* and < *TB_mortality_rate_smearpos_HIV1* | |
| *TB_mortality_rate_smearneg_HIV2* | Annual rate of mortality from smear- pulmonary TB for HIV+ART+ | Between 0.16 and 0.91 times *TB_mortality_rate_smearneg_HIV1*, and ≥ *TB_mortality_rate_smearneg_HIV0* | Dheda *et* al (2004)[33] and Lawn *et al* (2009)[34] |
| *TB_mortality_rate_treatment_DS* | Annual TB mortality rate when receiving TB treatment, for DS TB | ≥0 | Vary freely to fit data on treatment outcomes |
| *TB_mortality_rate_treatment_MDR* | Annual TB mortality rate when receiving TB treatment, for MDR TB | ≥0 | Vary freely to fit data on treatment outcomes |

| | | | |
|---|---|---|---|
| *TB_treatment_dropout_rate_DS* | Annual rate of dropping out of TB treatment for people with DS TB | ≥0 | Vary freely to fit data on treatment outcomes |
| *TB_treatment_dropout_rate_MDR* | Monthly rate of dropping out of TB treatment for people with MDR TB | ≥0 | Vary freely to fit data on treatment outcomes |
| *treatment_rate_HIV0_early* | Annual rate of starting treatment for HIV- people before *treatment_rate_change_year1* | ≥0 | Vary freely to fit data |
| *treatment_rate_HIV12_early* | Annual rate of starting treatment for HIV+ people before *treatment_rate_change_year1* | ≥0 | Vary freely to fit data |
| *treatment_rate_HIV0_late* | Annual rate of starting treatment for HIV- people from *treatment_rate_change_year1* | ≥0 | Vary freely to fit data |
| *treatment_rate_HIV12_late* | Annual rate of starting treatment for HIV+ people from *treatment_rate_change_year1* | ≥0 | Vary freely to fit data |
| *treatment_rate_change_year* | Year in which the values of the treatment start rate parameters change | 2010 | Data suggests treatment coverage was relatively stable from 2010[11] |
| *treatment_duration_DS* | Length of DS TB treatment in months | 6 | Managing TB in a New Era of Diagnostics[8] |
| *treatment_duration_MDR* | Length of MDR TB treatment in months | 24 until 2016, then 30% probability 24, 70% probability 11 | Expert opinion, WHO[7], and Managing TB in a New Era of Diagnostics[8] |
| *develop_tb_y1_rate_HIV0* | Annual rate of developing TB for HIV- people during the 1st year following infection, before | 0.0866 | Kasaie *et al*[35] |

| | | | |
|---|---|---|---|
| | *TB_parameter_change_year* | | |
| *develop_tb_y2_rate_HIV0* | Annual rate of developing TB for HIV- people during the 2nd year following infection, before *TB_parameter_change_year* | 0.0355 | Kasaie *et al*[35] |
| *develop_tb_y3_rate_HIV0* | Annual rate of developing TB for HIV- people during the 3rd year following infection, before *TB_parameter_change_year* | 0.0112 | Kasaie *et al*[35] |
| *develop_tb_y4_rate_HIV0* | Annual rate of developing TB for HIV- people during the 4th year following infection, before *TB_parameter_change_year* | $7.4 * 10^{-3}$ | Kasaie *et al*[35] |
| *develop_tb_y5_rate_HIV0* | Annual rate of developing TB for HIV- people during the 5th year following infection, before *B_parameter_change_year* | $2.4 * 10^{-3}$ | Kasaie *et al*[35] |
| *develop_tb_reactivation_rate_HIV0* | Annual rate of developing TB for HIV- people who have been infected for more than 5 years (late latent), before *TB_parameter_change_year* | $5.0 * 10^{-4}$ | Kasaie *et al*[35] |
| *develop_tb_y1_rate_HIV2* | Annual rate of developing TB for HIV+ART+ people during the 1st year following infection, before *TB_parameter_change_year* | 2 * *develop_tb_y1_ rate_HIV0* | Lawn *et al*[36] |
| *develop_tb_y2_rate_HIV2* | Annual rate of developing TB for HIV+ART+ people during the 2nd year following infection, before *TB_parameter_change_year* | 2 * *develop_tb_y2_ rate_HIV0* | Lawn *et al*[36] |

| | | | |
|---|---|---|---|
| *develop_tb_y3_rate_HIV2* | Annual rate of developing TB for HIV+ART+ people during the 3rd year following infection, before *TB_parameter_change_year* | 2 * *develop_tb_y3_rate_HIV0* | Lawn *et al*[36] |
| *develop_tb_y4_rate_HIV2* | Annual rate of developing TB for HIV+ART+ people during the 4th year following infection, before *TB_parameter_change_year* | 2 * *develop_tb_y4_rate_HIV0* | Lawn *et al*[36] |
| *develop_tb_y5_rate_HIV2* | Annual rate of developing TB for HIV+ART+ people during the 5th year following infection, before *TB_parameter_change_year* | 2 * *develop_tb_y5_rate_HIV0* | Lawn *et al*[36] |
| *develop_tb_reactivation_rate_HIV2* | Annual rate of developing TB for HIV+ART+ people who have been infected for more than 5 years (late latent), before *TB_parameter_change_year* | 2 * *develop_tb_reactivation_rate_HIV0* | Lawn *et al*[36] |
| *develop_tb_reactivation_rate_HIV1_early* | Annual rate of developing TB for HIV+ART- people who have been infected for more than 1 year, before *change_hiv1_parameters_year*, and before the adjustment that occurs from *TB_parameter_change_year* | > *develop_tb_reactivation_rate_HIV2* | |
| *develop_tb_reactivation_rate_HIV1_late* | Annual rate of developing TB for HIV+ART- people who have been infected for more than 1 year, from *change_hiv1_parameters_year* onwards, and before the adjustment that occurs from *TB_parameter_change_year* | > *develop_tb_reactivation_rate_HIV2* and < *develop_tb_reactivation_rate_HIV1_late* | |

| | | | |
|---|---|---|---|
| *increased_develop_tb_y1_rate_HIV1* | Increased rate of developing TB for HIV+ART- people, during the first year following infection compared to subsequent years | 5.14 | Dowdy and Chaisson (2009)[16] |
| *prop_smearpos_HIV0* | Proportion of HIV- people who develop TB, who develop smear+ disease | 0.45 | Corbett *et al* (2003)[37] |
| *prop_smearpos_HIV1* | Proportion of HIV positive people, not on ART, who develop TB, who develop smear+ disease | 0.35 | Corbett *et al* (2003)[37] |
| *prop_smearpos_HIV2* | Proportion of HIV positive people, on ART, who develop TB, who develop smear+ disease | 0.4 | Intermediate between HIV- and HIV+ART- |
| *introduce_MDR_year* | Year that MDR TB is introduced into the model | 2010 | Model population size large enough to prevent strain extinction |
| *seed_prop_MDR* | Proportion of people with Mtb infections who are seeded with MDR TB | 0.029 | Ismail *et al* 2018[38] |
| **HIV parameters** | | | |
| *HIV_intro_year* | Year that HIV is introduced into the model | 2000 | |
| *hiv_prev_initial_m0* | Proportion of males aged 15-29 seeded with HIV at its introduction in *HIV_intro_year* | 0.176 | 2002 HIV prevalence survey[39] |
| *hiv_prev_initial_m1* | Proportion of males aged 30-49 seeded with HIV at its introduction in *HIV_intro_year* | 0.177 | 2002 HIV prevalence survey[39] |
| *hiv_prev_initial_m2* | Proportion of males aged 50+ seeded with HIV at its introduction in *HIV_intro_year* | 0.073 | 2002 HIV prevalence survey[39] |

| | | | |
|---|---|---|---|
| *hiv_prev_initial_f0* | Proportion of males aged 15-29 seeded with HIV at its introduction in *HIV_intro_year* | 0.105 | 2002 HIV prevalence survey[39] |
| *hiv_prev_initial_f1* | Proportion of males aged 30-49 seeded with HIV at its introduction in *HIV_intro_year* | 0.174 | 2002 HIV prevalence survey[39] |
| *hiv_prev_initial_f2* | Proportion of males aged 50+ seeded with HIV at its introduction in *HIV_intro_year* | 0.064 | 2002 HIV prevalence survey[39] |
| *HIV1_mortality_rate* | Annual HIV mortality rate in HIV+ART- people | 0.1 | Mossong *et al* (2013)[40] |
| *HIV2_mortality_rate* | Annual HIV mortality rate in HIV+ART+ people | 0.0027 | Brinkhof *et al* (2009)[41] |
| *hiv_inc_early_f0* | Annual HIV incidence rate between *HIV_intro_year* and *HIV_inc_change_year1* in females aged 15-29 | 0-1 | Varied to fit data |
| *hiv_inc_early_f1* | Annual HIV incidence rate between *HIV_intro_year* and *HIV_inc_change_year1* in females aged 30-49 | 0-1 | Varied to fit data |
| *hiv_inc_early_f2* | Annual HIV incidence rate between *HIV_intro_year* and *HIV_inc_change_year1* in females aged 50-79 | 0-1 | Varied to fit data |
| *hiv_inc_early_m0* | Annual HIV incidence rate between *HIV_intro_year* and *HIV_inc_change_year1* in males aged 15-29 | 0-1 | Varied to fit data |
| *hiv_inc_early_m1* | Annual HIV incidence rate between *HIV_intro_year* and *HIV_inc_change_year1* in males aged 30-49 | 0-1 | Varied to fit data |
| *hiv_inc_early_m2* | Annual HIV incidence rate between *HIV_intro_year* and *HIV_inc_change_year1* in males aged 50-79 | 0-1 | Varied to fit data |

| | | | |
|---|---|---|---|
| *hiv_inc_mid_f0* | Annual HIV incidence rate between *HIV_inc_change_year1* and *HIV_inc_change_year2* in females aged 15-29 | 0-1 | Varied to fit data |
| *hiv_inc_mid_f1* | Annual HIV incidence rate between *HIV_inc_change_year1* and *HIV_inc_change_year2* in females aged 30-49 | 0-1 | Varied to fit data |
| *hiv_inc_mid_f2* | Annual HIV incidence rate between *HIV_inc_change_year1* and *HIV_inc_change_year2* in females aged 50+ | 0-1 | Varied to fit data |
| *hiv_inc_mid_m0* | Annual HIV incidence rate between *HIV_inc_change_year1* and *HIV_inc_change_year2* in males aged 15-29 | 0-1 | Varied to fit data |
| *hiv_inc_mid_m1* | Annual HIV incidence rate between *HIV_inc_change_year1* and *HIV_inc_change_year2* in males aged 30-49 | 0-1 | Varied to fit data |
| *hiv_inc_mid_m2* | Annual HIV incidence rate between *HIV_inc_change_year1* and *HIV_inc_change_year2* in males aged 50+ | 0-1 | Varied to fit data |
| *HIV_inc_reduction_late_m* | Annual relative change in HIV incidence in males from *HIV_inc_change_year2*, compared to the incidence in the same age group between *HIV_inc_change_year1* and *HIV_inc_change_year2* | 0-1 | Varied to fit data |

| | | | |
|---|---|---|---|
| *HIV_inc_reduction_late_f* | Annual relative change in HIV incidence in females from *HIV_inc_change_year2*, compared to the incidence in the same age group between *HIV_inc_change_year1* and *HIV_inc_change_year2* | 0-1 | Varied to fit data |
| *HIV_inc_change_year1* | Year at which HIV incidence parameters change for the first time | 2012 | Estimated year at which HIV incidence started to decline in the DSA area[17] |
| *HIV_inc_change_year2* | Year at which HIV incidence parameters change for the second time | 2021 | To allow projected future trend in HIV prevalence to be simulated |
| *ART_intro_year* | Year that ART is introduced into the model | 2005 | Coverage of ART was very low in South Africa prior to 2005[42] |
| *ART_start_rate_change_year* | Year at which the rate of starting ART changes | 2013 | Changed year after first ART prevalence fitting target |
| *ART_start_rate_early_m* | Annual rate of starting ART for HIV+ males between *ART_intro_year* and *ART_start_rate_change_year* | 0-1 | Varied to fit data |
| *ART_start_rate_early_f* | Annual rate of starting ART for HIV+ females between *ART_intro_year* and *ART_start_rate_change_year* | 0-1 | Varied to fit data |

| | | | |
|---|---|---|---|
| *ART_start_rate_late_m* | Annual rate of starting ART for HIV+ males after *ART_start_rate_change_year* | 0-1 | Varied to fit data |
| *ART_start_rate_late_f* | Annual rate of starting ART for HIV+ females after *ART_start_rate_change_year* | 0-1 | Varied to fit data |
| *change_HIV1_parameters_year* | Year at which the values of *reinfection_relative_risk_HIV1* and *develop_tb_reactivation_rate_HIV1* are changed from their 'early' values to their 'late' values | >2005 | After the introduction of ART in the model |
| **Demography parameters** | | | |
| *initial_pop_size* | Initial population size | 10000 | Balance of model run times and degree of stochasticity in individual runs |
| *initial_m_age0* | Initial proportion of males in the age group 15-29 | 0.432 | Same as the desired age distribution in 2018 |
| *initial_m_age1* | Initial proportion of males in the age group 30-49 | 0.387 | Same as the desired age distribution in 2018 |
| *initial_m_age2* | Initial proportion of males in the age group 50-79 | 0.181 | Same as the desired age distribution in 2018 |
| *initial_f_age0* | Initial proportion of females in the age group 15-29 | 0.382 | Same as the desired age distribution in 2018 |
| *initial_f_age1* | Initial proportion of females in the age group 30-49 | 0.363 | Same as the desired age distribution in 2018 |

| | | | |
|---|---|---|---|
| *initial_f_age2* | Initial proportion of females in the age group 50-79 | 0.255 | Same as the desired age distribution in 2018 |
| *birth_rate* | Annual birth rate per person | 0-1 | Varied to fit data |
| *mean_hh_size* | Mean simulated household size (individuals aged 15+ years) | 3.64 | Estimated from empirical data (see section 'Household sizes') |
| *hhsize_parameter_a* | See section 'Household size' | 0.2 | Estimated from empirical data (see section 'Household sizes') |
| *hhsize_parameter_b* | See section 'Household size' | 4.2 | Estimated from empirical data (see section 'Household sizes') |
| *mortality_rate_m_age0* | Annual baseline (non-TB or HIV) mortality for males aged 15-29 | 0-1 | Varied to fit data |
| *mortality_rate_m_age1* | Annual baseline (non-TB or HIV) mortality for males aged 30-49 | 0-1 | Varied to fit data |
| *mortality_rate_m_age2* | Annual baseline (non-TB or HIV) mortality for males aged 50+ | 0-1 | Varied to fit data |

| | | | |
|---|---|---|---|
| *mortality_rate_f_age0* | Annual baseline (non-TB or HIV) mortality for females aged 15-29 | 0-1 | Varied to fit data |
| *mortality_rate_f_age1* | Annual baseline (non-TB or HIV) mortality for females aged 30-49 | 0-1 | Varied to fit data |
| *mortality_rate_f_age2* | Annual baseline (non-TB or HIV) mortality for females aged 50+ | 0-1 | Varied to fit data |
| **Contact time parameters** | | | |
| *contact_time_each_hh_mem* | Minutes of indoor contact time per month between each household member | 34328 | Social contact survey |
| *contact_time_other_m_age0_HIV01* | Minutes of contact time per month in other settings for HIV- males and HIV+ART- males, aged 15-29 | 138917 | Social contact survey |
| *contact_time_other_m_age0_HIV2* | Minutes of contact time per month in other settings for HIV+ART+ males, aged 15-29 | 116328 | Social contact survey |
| *contact_time_other_m_age1_HIV01* | Minutes of contact time per month in other settings for HIV- males and HIV+ART- males, aged 30-49 | 98160 | Social contact survey |
| *contact_time_other_m_age1_HIV2* | Minutes of contact time per month in other settings for HIV+ART+ males, aged 30-49 | 75571 | Social contact survey |
| *contact_time_other_m_age2_HIV01* | Minutes of contact time per month in other settings for HIV- males and HIV+ART- males, aged 50+ | 94046 | Social contact survey |
| *contact_time_other_m_age2_HIV2* | Minutes of contact time per month in other settings for HIV+ART+ males, aged 50+ | 71457 | Social contact survey |

| | | | |
|---|---|---|---|
| *contact_time_other_f_age0_HIV01* | Minutes of contact time per month in other settings for HIV- females and HIV+ART- females, aged 15-29 | 143625 | Social contact survey |
| *contact_time_other_f_age0_HIV2* | Minutes of contact time per month in other settings for HIV+ART+ females, aged 15-29 | 121036 | Social contact survey |
| *contact_time_other_f_age1_HIV01* | Minutes of contact time per month in other settings for HIV- females and HIV+ART- females, aged 30-49 | 102867 | Social contact survey |
| *contact_time_other_f_age1_HIV2* | Minutes of contact time per month in other settings for HIV+ART+ females, aged 30-49 | 80278 | Social contact survey |
| *contact_time_other_f_age2_HIV01* | Minutes of contact time per month in other settings for HIV- females and HIV+ART- females, aged 50+ | 98754 | Social contact survey |
| *contact_time_other_f_age2_HIV2* | Minutes of contact time per month in other settings for HIV+ART+ females, aged 50+ | 76164 | Social contact survey |
| *contact_time_clinic_m_HIV0_low* | Minutes of contact time per month in clinics for HIV- males, in the low clinic visiting group | 493 (varied in sensitivity analyses) | Social contact survey |
| *contact_time_clinic_m_HIV1_low* | Minutes of contact time per month in clinics for HIV+ART- males, in the low clinic visiting group | 493 (varied in sensitivity analyses) | Social contact survey |
| *contact_time_clinic_m_HIV2_low* | Minutes of contact time per month in clinics for HIV+ART+ males, in the low clinic visiting group | 3468 (varied in sensitivity analyses) | Social contact survey |
| *contact_time_clinic_f_HIV0_low* | Minutes of contact time per month in clinics for HIV- females, in the low clinic visiting group | 2322 (varied in sensitivity analyses) | Social contact survey |
| *contact_time_clinic_f_HIV1_low* | Minutes of contact time per month in clinics for HIV+ART- females, in the low clinic visiting group | 2322 (varied in sensitivity analyses) | Social contact survey |

| | | | |
|---|---|---|---|
| *contact_time_clinic_f_HIV2_low* | Minutes of contact time per month in clinics for HIV+ART+ females, in the low clinic visiting group | 8276 (varied in sensitivity analyses) | Social contact survey |
| *contact_time_clinic_m_HIV0_high* | Minutes of contact time per month in clinics for HIV- males, in the high clinic visiting group | 5507 (varied in sensitivity analyses) | Social contact survey |
| *contact_time_clinic_m_HIV1_high* | Minutes of contact time per month in clinics for HIV+ART- males, in the high clinic visiting group | 5507 (varied in sensitivity analyses) | Social contact survey |
| *contact_time_clinic_m_HIV2_high* | Minutes of contact time per month in clinics for HIV+ART+ males, in the high clinic visiting group | 8400 (varied in sensitivity analyses) | Social contact survey |
| *contact_time_clinic_f_HIV0_high* | Minutes of contact time per month in clinics for HIV- females, in the high clinic visiting group | 8609 (varied in sensitivity analyses) | Social contact survey |
| *contact_time_clinic_f_HIV1_high* | Minutes of contact time per month in clinics for HIV+ART- females, in the high clinic visiting group | 8609 (varied in sensitivity analyses) | Social contact survey |
| *contact_time_clinic_f_HIV2_high* | Minutes of contact time per month in clinics for HIV+ART+ females, in the high clinic visiting group | 8276 (varied in sensitivity analyses) | Social contact survey |
| *increased_contact_time_clinics_tb* | Increased contact time in clinics for people with TB compared to people without | >1 | Varied freely, to fit data |
| *clinic_rate_switch_prob* | Probability of switching clinic visiting group every six months | 0.25 (varied in sensitivity analysis) | Plausible value. Effects explored in sensitivity analysis |
| *ventilation_weight_home* | Modifier of *transmission_prob* for contact time between household members, incorporating effects of different mean ventilation rates by location type | 2.8 | Lygizos *et al* 2013[4] and Beckwith *et al*[5] |

| | | | |
|---|---|---|---|
| *ventilation_weight_clinic* | Modifier of *transmission_prob* for contact time in clinics, incorporating effects of different mean ventilation rates by location type | 1 (varied in sensitivity analysis) | |
| *ventilation_weight_other* | Modifier of *transmission_prob* for contact time in other locations, incorporating effects of different mean ventilation rates by location type | 1 | Taylor *et al* 2016[6] and Beckwith *et al*[5] |
| **Intervention parameters** | | | |
| *int_RR_trans_clinics* | Modifier of risk of infection per minute contact occurring in clinics | 1 until 2021, then value dependent on simulated intervention | See section 'Interventions' |
| *int_RR_contact_clinics_HIV01* | Modifier of mean contact hours in clinics for people who are HIV- or HIV+ART- | 1 until 2021, then value dependent on simulated intervention | See section 'Interventions' |
| *int_RR_contact_clinics_HIV2* | Modifier of mean contact hours in clinics for people who are HIV+ART+ | 1 until 2021, then value dependent on simulated intervention | See section 'Interventions' |

**Table S9. Description of model input parameters, values or plausible ranges, and data sources.**

#### 2.16 Model fitting targets

| Description | Calibration target/Plausible range | Source |
| --- | --- | --- |
| Growth in population size between 2015 and 2019 | 3.4% | Mid-year population estimates 2019[43] |
| Proportion of the population who are male in 2018 | 48% | Mid-year population estimates 2018[44] |
| Proportion of simulated men aged 15-29 | 43% | Mid-year population estimates 2018[44] |
| Proportion of simulated men aged 30-49 | 39% | Mid-year population estimates 2018[44] |
| Proportion of simulated men aged 50+ | 18% | Mid-year population estimates 2018[44] |
| Proportion of simulated women aged 15-29 | 38% | Mid-year population estimates 2018[44] |
| Proportion of simulated women aged 30-49 | 36% | Mid-year population estimates 2018[44] |
| Proportion of simulated women aged 50+ | 25% | Mid-year population estimates 2018[44] |
| HIV prevalence in men aged 15-29, in 2011 | 7% | Vandormael *et al* (2019)[17] |
| HIV prevalence in men aged 30-49, in 2011 | 48% | Vandormael *et al* (2019)[17] |
| HIV prevalence in women aged 15-29, in 2011 | 26% | Vandormael *et al* (2019)[17] |
| HIV prevalence in women aged 30-49, in 2011 | 48% | Vandormael *et al* (2019)[17] |
| HIV prevalence in men aged 15-29, in 2017 | 8% | Vandormael *et al* (2019)[17] |
| HIV prevalence in men aged 30-49, in 2017 | 44% | Vandormael *et al* (2019)[17] |
| HIV prevalence in men aged 50+, in 2017 | 30% | Vandormael *et al* (2019)[17] |
| HIV prevalence in women aged 15-29, in 2017 | 25% | Vandormael *et al* (2019)[17] |
| HIV prevalence in women aged 30-49, in 2017 | 59% | Vandormael *et al* (2019)[17] |

| | | |
|---|---|---|
| HIV prevalence in women aged 50+, in 2017 | 35% | Vandormael *et al* (2019)[17] |
| Proportion of HIV positive people on ART in 2012 | 25-45% | Vandormael *et al* (2019)[17] |
| Proportion of HIV positive people aged 15-29 on ART in 2017 | 49% | Vandormael *et al* (2019)[17] |
| Proportion of HIV positive people aged 30-49 on ART in 2017 | 74% | Vandormael *et al* (2019)[17] |
| Proportion of HIV positive people aged 50+ on ART in 2017 | 86% | Vandormael *et al* (2019)[17] |
| Proportion of HIV positive men on ART in 2017 | 63% | Vandormael *et al* (2019)[17] |
| Proportion of HIV positive women on ART in 2017 | 73% | Vandormael *et al* (2019)[17] |
| Annual incidence of TB per 100,000 population in 2011 | 1433 (1107-1803) | Notification data for KZN[28], adjusted for under-reporting, assuming that the proportion of cases notified is the same for KZN as for South Africa as a whole[11] |
| Annual incidence of TB per 100,000 population in 2018 | 658 (472-874) | Notification data for KZN[28], adjusted for under-reporting, assuming that the proportion of cases notified is the same for KZN as for South Africa as a whole[11] |
| Proportion of incident TB that is in HIV positive people in 2018 | 0.58 | Data on patients starting TB treatment in KZN[28], assuming that the proportion of incident TB that is in HIV positive people is the same as the proportion of people starting TB treatment who are HIV positive (as is assumed by WHO for South Africa as a whole[11]). |
| Proportion of incident TB that is MDR in 2012 | 0.029 | Ismail *et al* 2018[38] |
| Proportion of incident TB that is MDR in 2018 | 0.031 | Estimated proportion of TB cases starting treatment in 2018 who have MDR TB. Unpublished, provisional data from the National Institute for Communicable Diseases |

| | | |
|---|---|---|
| Annual HIV negative TB mortality rate per 100,000 population in 2018 | 47 (34-63) | Calculated from estimated incidence in HIV- people in KZN in 2018, and estimated case fatality ratio for TB in HIV- people in South Africa[11] |
| Annual HIV positive TB mortality rate per 100,000 population in 2018 | 92 (66-122) | Calculated from estimated incidence in HIV positive people in KZN in 2018, and estimated case fatality ratio for TB in HIV positive people in South Africa[11] |
| Proportion of people starting TB treatment who are HIV positive in 2018 | 0.58 | Data on patients starting TB treatment in KZN[28] |
| Ratio of cases starting treatment to estimated incidence in 2000 | 57% (40%-89%) | WHO global TB report 2019[11] |
| Ratio of cases starting treatment to estimated incidence in 2018 | 76% (57%-100%) | WHO global TB report 2019[11] |
| Proportion starting treatment in 2017 who complete treatment, DS TB | 78% | WHO global TB report 2019[11] |
| Proportion starting treatment in 2017 who complete treatment, MDR TB | 54% | WHO global TB report 2019[11] |
| Proportion starting treatment in 2017 who die while on treatment, DS TB | 11% | Data from KZN[28] |
| Proportion starting treatment in 2017 who die while on treatment, MDR TB | 23% | Data from KZN[28] |
| Proportion starting treatment in 2017 who dropped out of treatment, DS TB | 11% | Data from KZN[28] |

| | | |
|---|---|---|
| Proportion starting treatment in 2017 who dropped out of treatment, MDR TB | 23% | Data from KZN[28] |
| Increased prevalence of TB in clinic attendees, compared to the general population, in 2019 | 1.86 | Govender *et al* (2020)[3] |
| Proportion of incident TB that results from transmission between household members, in 2018 | 13.5% | McCreesh and White (2018)[14] |
| Relative change in HIV prevalence in men between 2020 and 2030 | -16.2% | Estimates from Thembisa model[18 19] |
| Relative change in HIV prevalence in women between 2020 and 2030 | -5.7% | Estimates from Thembisa model[18 19] |
| Relative change in ART coverage among HIV+ men between 2020 and 2030 | 5.4% | Estimates from Thembisa model[18 19] |
| Relative change in ART coverage among HIV+ women between 2020 and 2030 | 2.0% | Estimates from Thembisa model[18 19] |

**Table S10. Model fitting targets in the best estimate scenario.** Where no ranges are given, fits were considered acceptable if they were within ±20% of the target value.

### 3 Model results

#### 3.1 Calibrated input parameter values

| | Best estimate | Proportion of outside-household contact time occurring in clinics | | Proportion of disease from household transmission | | Ventilation rates in clinics | | Prevalence of TB in clinic attendees | | Movement between high and low clinic visiting groups | | HIV+ART clinic visiting | | Future HIV incidence | |
|---|---|---|---|---|---|---|---|---|---|---|---|---|---|---|---|
| | | Low | High | Low | High | Low | High | Low | High | Low | High | Low | High | Low | High |
| *birth_rate* | 0.0021 | 0.0021 | 0.0021 | 0.0021 | 0.0021 | 0.0021 | 0.0021 | 0.0021 | 0.0021 | 0.0021 | 0.0021 | 0.0021 | 0.0021 | 0.0021 | 0.0021 |
| *mortality_rate_m_age0* | 0 | 0 | 0 | 0 | 0 | 0 | 0 | 0 | 0 | 0 | 0 | 0 | 0 | 0 | 0 |
| *mortality_rate_m_age1* | 0 | 0 | 0 | 0 | 0 | 0 | 0 | 0 | 0 | 0 | 0 | 0 | 0 | 0 | 0 |
| *mortality_rate_m_age2* | 0.055 | 0.055 | 0.055 | 0.055 | 0.055 | 0.055 | 0.055 | 0.055 | 0.055 | 0.055 | 0.055 | 0.055 | 0.055 | 0.055 | 0.055 |
| *mortality_rate_f_age0* | 0 | 0 | 0 | 0 | 0 | 0 | 0 | 0 | 0 | 0 | 0 | 0 | 0 | 0 | 0 |
| *mortality_rate_f_age1* | 0 | 0 | 0 | 0 | 0 | 0 | 0 | 0 | 0 | 0 | 0 | 0 | 0 | 0 | 0 |
| *mortality_rate_f_age2* | 0.025 | 0.025 | 0.025 | 0.025 | 0.025 | 0.025 | 0.025 | 0.025 | 0.025 | 0.025 | 0.025 | 0.025 | 0.025 | 0.025 | 0.025 |
| ***transmission_prob_early*** | 1.11E-05 | 1.16E-05 | 1.09E-05 | 1.03E-05 | 1.18E-05 | 1.13E-05 | 1.04E-05 | 1.13E-05 | 1.06E-05 | 1.11E-05 | 1.11E-05 | 1.11E-05 | 1.11E-05 | 1.11E-05 | 1.10E-05 |
| *TB_parameter_change_year* | 2014 | 2014 | 2014 | 2014 | 2014 | 2014 | 2014 | 2014 | 2014 | 2014 | 2014 | 2014 | 2014 | 2014 | 2014 |
| *decreased_tb_rates_late* | 0.86 | 0.86 | 0.86 | 0.86 | 0.86 | 0.86 | 0.86 | 0.86 | 0.86 | 0.86 | 0.86 | 0.86 | 0.86 | 0.86 | 0.86 |
| *treatment_rate_HIV0_early* | 0.48 | 0.48 | 0.48 | 0.48 | 0.48 | 0.48 | 0.48 | 0.48 | 0.48 | 0.48 | 0.48 | 0.48 | 0.48 | 0.48 | 0.48 |
| *treatment_rate_HIV12_early* | 0.78 | 0.78 | 0.78 | 0.78 | 0.78 | 0.78 | 0.78 | 0.78 | 0.78 | 0.78 | 0.78 | 0.78 | 0.78 | 0.78 | 0.78 |
| *treatment_rate_HIV0_late* | 0.68 | 0.68 | 0.68 | 0.68 | 0.68 | 0.68 | 0.68 | 0.68 | 0.68 | 0.68 | 0.68 | 0.68 | 0.68 | 0.68 | 0.68 |
| *treatment_rate_HIV12_late* | 0.81 | 0.81 | 0.81 | 0.81 | 0.81 | 0.81 | 0.81 | 0.81 | 0.81 | 0.81 | 0.81 | 0.81 | 0.81 | 0.81 | 0.81 |
| *treatment_rate_HIV0_late* | 0.68 | 0.68 | 0.68 | 0.68 | 0.68 | 0.68 | 0.68 | 0.68 | 0.68 | 0.68 | 0.68 | 0.68 | 0.68 | 0.68 | 0.68 |
| *treatment_rate_HIV12_late* | 0.81 | 0.81 | 0.81 | 0.81 | 0.81 | 0.81 | 0.81 | 0.81 | 0.81 | 0.81 | 0.81 | 0.81 | 0.81 | 0.81 | 0.81 |
| *TB_mortality_rate_smearneg_HIV0* | 0.025 | 0.025 | 0.025 | 0.025 | 0.025 | 0.025 | 0.025 | 0.025 | 0.025 | 0.025 | 0.025 | 0.025 | 0.025 | 0.025 | 0.025 |
| *TB_mortality_rate_smearneg_HIV1* | 0.60 | 0.60 | 0.60 | 0.60 | 0.60 | 0.60 | 0.60 | 0.60 | 0.60 | 0.60 | 0.60 | 0.60 | 0.60 | 0.60 | 0.60 |
| *TB_mortality_rate_smearneg_HIV2* | 0.10 | 0.10 | 0.10 | 0.10 | 0.10 | 0.10 | 0.10 | 0.10 | 0.10 | 0.10 | 0.10 | 0.10 | 0.10 | 0.10 | 0.10 |
| *TB_mortality_rate_smearpos_HIV0* | 0.39 | 0.39 | 0.39 | 0.39 | 0.39 | 0.39 | 0.39 | 0.39 | 0.39 | 0.39 | 0.39 | 0.39 | 0.39 | 0.39 | 0.39 |

| | | | | | | | | | | | | | | | |
|---|---|---|---|---|---|---|---|---|---|---|---|---|---|---|---|
| TB_mortality_rate_smearpos_HIV1 | 0.80 | 0.80 | 0.80 | 0.80 | 0.80 | 0.80 | 0.80 | 0.80 | 0.80 | 0.80 | 0.80 | 0.80 | 0.80 | 0.80 | 0.80 |
| TB_mortality_rate_smearpos_HIV2 | 0.39 | 0.39 | 0.39 | 0.39 | 0.39 | 0.39 | 0.39 | 0.39 | 0.39 | 0.39 | 0.39 | 0.39 | 0.39 | 0.39 | 0.39 |
| TB_mortality_rate_treatment_DS | 0.12 | 0.12 | 0.12 | 0.12 | 0.12 | 0.12 | 0.12 | 0.12 | 0.12 | 0.12 | 0.12 | 0.12 | 0.12 | 0.12 | 0.12 |
| TB_mortality_rate_treatment_MDR | 0.14 | 0.14 | 0.14 | 0.14 | 0.14 | 0.14 | 0.14 | 0.14 | 0.14 | 0.14 | 0.14 | 0.14 | 0.14 | 0.14 | 0.14 |
| TB_treatment_dropout_rate_DS | 0.011 | 0.011 | 0.011 | 0.011 | 0.011 | 0.011 | 0.011 | 0.011 | 0.011 | 0.011 | 0.011 | 0.011 | 0.011 | 0.011 | 0.011 |
| TB_treatment_dropout_rate_MDR | 0.011 | 0.011 | 0.011 | 0.011 | 0.011 | 0.011 | 0.011 | 0.011 | 0.011 | 0.011 | 0.011 | 0.011 | 0.011 | 0.011 | 0.011 |
| develop_tb_reactivation_rate_HIV1_early | 0.20 | 0.20 | 0.20 | 0.20 | 0.20 | 0.20 | 0.20 | 0.20 | 0.20 | 0.20 | 0.20 | 0.20 | 0.20 | 0.20 | 0.20 |
| develop_tb_reactivation_rate_HIV1_late | 0.080 | 0.080 | 0.080 | 0.080 | 0.080 | 0.080 | 0.080 | 0.080 | 0.080 | 0.080 | 0.080 | 0.080 | 0.080 | 0.080 | 0.080 |
| reinfection_relative_risk_HIV1_early | 0.90 | 0.90 | 0.90 | 0.90 | 0.90 | 0.90 | 0.90 | 0.90 | 0.90 | 0.90 | 0.90 | 0.90 | 0.90 | 0.90 | 0.90 |
| reinfection_relative_risk_HIV1_late | 0.60 | 0.60 | 0.60 | 0.60 | 0.60 | 0.60 | 0.60 | 0.60 | 0.60 | 0.60 | 0.60 | 0.60 | 0.60 | 0.60 | 0.60 |
| reinfection_relative_risk_HIV2 | 0.50 | 0.50 | 0.50 | 0.50 | 0.50 | 0.50 | 0.50 | 0.50 | 0.50 | 0.50 | 0.50 | 0.50 | 0.50 | 0.50 | 0.50 |
| change_HIV0_parameters_year | 2007 | 2007 | 2007 | 2007 | 2007 | 2007 | 2007 | 2007 | 2007 | 2007 | 2007 | 2007 | 2007 | 2007 | 2007 |
| hiv_inc_early_f0 | 0.066 | 0.066 | 0.066 | 0.066 | 0.066 | 0.066 | 0.066 | 0.066 | 0.066 | 0.066 | 0.066 | 0.066 | 0.066 | 0.066 | 0.066 |
| hiv_inc_early_f1 | 0.081 | 0.081 | 0.081 | 0.081 | 0.081 | 0.081 | 0.081 | 0.081 | 0.081 | 0.081 | 0.081 | 0.081 | 0.081 | 0.081 | 0.081 |
| hiv_inc_early_f2 | 0.023 | 0.023 | 0.023 | 0.023 | 0.023 | 0.023 | 0.023 | 0.023 | 0.023 | 0.023 | 0.023 | 0.023 | 0.023 | 0.023 | 0.023 |
| hiv_inc_early_m0 | 0.015 | 0.015 | 0.015 | 0.015 | 0.015 | 0.015 | 0.015 | 0.015 | 0.015 | 0.015 | 0.015 | 0.015 | 0.015 | 0.015 | 0.015 |
| hiv_inc_early_m1 | 0.12 | 0.12 | 0.12 | 0.12 | 0.12 | 0.12 | 0.12 | 0.12 | 0.12 | 0.12 | 0.12 | 0.12 | 0.12 | 0.12 | 0.12 |
| hiv_inc_early_m2 | 0 | 0 | 0 | 0 | 0 | 0 | 0 | 0 | 0 | 0 | 0 | 0 | 0 | 0 | 0 |
| hiv_inc_mid_f0 | 0.055 | 0.055 | 0.055 | 0.055 | 0.055 | 0.055 | 0.055 | 0.055 | 0.055 | 0.055 | 0.055 | 0.055 | 0.055 | 0.055 | 0.055 |
| hiv_inc_mid_f1 | 0.06 | 0.06 | 0.06 | 0.06 | 0.06 | 0.06 | 0.06 | 0.06 | 0.06 | 0.06 | 0.06 | 0.06 | 0.06 | 0.06 | 0.06 |
| hiv_inc_mid_f2 | 0.023 | 0.023 | 0.023 | 0.023 | 0.023 | 0.023 | 0.023 | 0.023 | 0.023 | 0.023 | 0.023 | 0.023 | 0.023 | 0.023 | 0.023 |
| hiv_inc_mid_m0 | 0.017 | 0.017 | 0.017 | 0.017 | 0.017 | 0.017 | 0.017 | 0.017 | 0.017 | 0.017 | 0.017 | 0.017 | 0.017 | 0.017 | 0.017 |
| hiv_inc_mid_m1 | 0.071 | 0.071 | 0.071 | 0.071 | 0.071 | 0.071 | 0.071 | 0.071 | 0.071 | 0.071 | 0.071 | 0.071 | 0.071 | 0.071 | 0.071 |
| hiv_inc_mid_m2 | 0 | 0 | 0 | 0 | 0 | 0 | 0 | 0 | 0 | 0 | 0 | 0 | 0 | 0 | 0 |
| **HIV_inc_reduction_late_m** | 0.46 | 0.46 | 0.46 | 0.46 | 0.46 | 0.46 | 0.46 | 0.46 | 0.46 | 0.46 | 0.46 | 0.46 | 0.46 | 0.23 | 0.73 |
| **HIV_inc_reduction_late_f** | 0.32 | 0.32 | 0.32 | 0.32 | 0.32 | 0.32 | 0.32 | 0.32 | 0.32 | 0.32 | 0.32 | 0.32 | 0.32 | 0.16 | 0.66 |
| ART_start_rate_early_m | 0.032 | 0.032 | 0.032 | 0.032 | 0.032 | 0.032 | 0.032 | 0.032 | 0.032 | 0.032 | 0.032 | 0.032 | 0.032 | 0.032 | 0.032 |
| ART_start_rate_early_f | 0.048 | 0.048 | 0.048 | 0.048 | 0.048 | 0.048 | 0.048 | 0.048 | 0.048 | 0.048 | 0.048 | 0.048 | 0.048 | 0.048 | 0.048 |

| | | | | | | | | | | | | | | | |
|---|---|---|---|---|---|---|---|---|---|---|---|---|---|---|---|
| *ART_start_rate_late_m* | 0.16 | 0.16 | 0.16 | 0.16 | 0.16 | 0.16 | 0.16 | 0.16 | 0.16 | 0.16 | 0.16 | 0.16 | 0.16 | 0.16 | 0.16 |
| *ART_start_rate_late_f* | 0.22 | 0.22 | 0.22 | 0.22 | 0.22 | 0.22 | 0.22 | 0.22 | 0.22 | 0.22 | 0.22 | 0.22 | 0.22 | 0.22 | 0.22 |
| ***increased_contact_time_clinics_tb*** | 1.42 | 1.42 | 1.42 | 1.42 | 1.42 | 1.42 | 1.42 | 1.00 | 2.42 | 1.42 | 1.42 | 1.42 | 1.42 | 1.42 | 1.42 |
| ***infectiousness_var*** | 33 | 35 | 33 | 18 | 72 | 33 | 33 | 33 | 33 | 33 | 33 | 33 | 33 | 33 | 33 |

**Table S11. Fitted input parameter values in the best estimate scenario and sensitivity analysis scenarios.** 'Low' and 'high' refer to changes that decrease and increase the proportion of disease that results from transmission in clinics respectively. Parameter names are given in bold if the fitted value varied between scenarios

#### 3.2 Fit to data

| | Target (best estimate scenario) | Best estimate | Proportion of outside-household contact time occurring in clinics | | Proportion of disease from household transmission | | Ventilation rates in clinics | | Prevalence of TB in clinic attendees | | Movement between high and low clinic visiting groups | | HIV+ART- clinic visiting | | Future HIV incidence | |
|---|---|---|---|---|---|---|---|---|---|---|---|---|---|---|---|---|
| | | | Low | High | Low | High | Low | High | Low | High | Low | High | Low | High | Low | High |
| Growth in population size between 2015 and 2019 | 0.034 | 0.033 | 0.033 | 0.034 | 0.033 | 0.033 | 0.033 | 0.033 | 0.033 | 0.033 | 0.033 | 0.033 | 0.033 | 0.034 | 0.033 | 0.033 |
| Proportion of the population who are male in 2018 | 0.48 | 0.50 | 0.50 | 0.50 | 0.50 | 0.50 | 0.50 | 0.50 | 0.50 | 0.50 | 0.50 | 0.50 | 0.50 | 0.50 | 0.50 | 0.50 |
| Proportion of simulated men aged 15-29 | 0.43 | 0.46 | 0.46 | 0.46 | 0.46 | 0.46 | 0.46 | 0.46 | 0.46 | 0.46 | 0.46 | 0.46 | 0.46 | 0.46 | 0.46 | 0.46 |
| Proportion of simulated men aged 30-49 | 0.39 | 0.39 | 0.39 | 0.39 | 0.39 | 0.39 | 0.39 | 0.39 | 0.39 | 0.39 | 0.39 | 0.39 | 0.39 | 0.39 | 0.39 | 0.39 |
| Proportion of simulated men aged 50+ | 0.18 | 0.15 | 0.15 | 0.15 | 0.15 | 0.15 | 0.15 | 0.15 | 0.15 | 0.15 | 0.15 | 0.15 | 0.15 | 0.15 | 0.15 | 0.15 |
| Proportion of simulated women aged 15-29 | 0.38 | 0.43 | 0.43 | 0.43 | 0.43 | 0.43 | 0.43 | 0.43 | 0.43 | 0.43 | 0.43 | 0.43 | 0.43 | 0.43 | 0.43 | 0.43 |
| Proportion of simulated women aged 30-49 | 0.36 | 0.33 | 0.33 | 0.33 | 0.33 | 0.33 | 0.33 | 0.33 | 0.33 | 0.33 | 0.33 | 0.33 | 0.33 | 0.33 | 0.33 | 0.33 |
| Proportion of simulated women aged 50+ | 0.25 | 0.24 | 0.24 | 0.24 | 0.24 | 0.24 | 0.24 | 0.24 | 0.24 | 0.24 | 0.24 | 0.24 | 0.24 | 0.24 | 0.24 | 0.24 |
| HIV prevalence in men aged 15-29, in 2011 | 0.070 | 0.074 | 0.073 | 0.073 | 0.07 | 0.073 | 0.074 | 0.074 | 0.073 | 0.074 | 0.074 | 0.074 | 0.074 | 0.073 | 0.074 | 0.074 |
| HIV prevalence in men aged 30-49, in 2011 | 0.48 | 0.50 | 0.50 | 0.50 | 0.50 | 0.50 | 0.50 | 0.50 | 0.50 | 0.50 | 0.50 | 0.50 | 0.50 | 0.50 | 0.50 | 0.50 |
| HIV prevalence in women aged 15-29, in 2011 | 0.26 | 0.27 | 0.27 | 0.27 | 0.27 | 0.27 | 0.27 | 0.27 | 0.27 | 0.27 | 0.27 | 0.27 | 0.27 | 0.27 | 0.27 | 0.27 |

| | | | | | | | | | | | | | | | | |
|---|---|---|---|---|---|---|---|---|---|---|---|---|---|---|---|---|
| HIV prevalence in women aged 30-49, in 2011 | 0.48 | 0.50 | 0.50 | 0.49 | 0.50 | 0.50 | 0.50 | 0.50 | 0.50 | 0.50 | 0.50 | 0.50 | 0.50 | 0.49 | 0.50 | 0.50 |
| HIV prevalence in men aged 15-29, in 2017 | 0.080 | 0.083 | 0.083 | 0.083 | 0.08 | 0.083 | 0.083 | 0.083 | 0.083 | 0.083 | 0.083 | 0.083 | 0.083 | 0.083 | 0.083 | 0.083 |
| HIV prevalence in men aged 30-49, in 2017 | 0.44 | 0.45 | 0.45 | 0.45 | 0.45 | 0.45 | 0.45 | 0.45 | 0.45 | 0.45 | 0.45 | 0.45 | 0.45 | 0.45 | 0.45 | 0.45 |
| HIV prevalence in men aged 50+, in 2017 | 0.30 | 0.34 | 0.34 | 0.34 | 0.34 | 0.34 | 0.34 | 0.34 | 0.34 | 0.34 | 0.34 | 0.34 | 0.34 | 0.34 | 0.34 | 0.34 |
| HIV prevalence in women aged 15-29, in 2017 | 0.25 | 0.26 | 0.26 | 0.26 | 0.26 | 0.26 | 0.26 | 0.26 | 0.26 | 0.26 | 0.26 | 0.26 | 0.26 | 0.26 | 0.26 | 0.26 |
| HIV prevalence in women aged 30-49, in 2017 | 0.59 | 0.56 | 0.56 | 0.56 | 0.57 | 0.56 | 0.56 | 0.56 | 0.56 | 0.56 | 0.56 | 0.56 | 0.57 | 0.56 | 0.56 | 0.57 |
| HIV prevalence in women aged 50+, in 2017 | 0.35 | 0.39 | 0.39 | 0.39 | 0.39 | 0.39 | 0.39 | 0.39 | 0.39 | 0.39 | 0.39 | 0.39 | 0.39 | 0.39 | 0.39 | 0.39 |
| Proportion of HIV positive people on ART in 2012 | 25-45% | 0.32 | 0.32 | 0.32 | 0.32 | 0.32 | 0.32 | 0.32 | 0.32 | 0.32 | 0.32 | 0.32 | 0.32 | 0.32 | 0.32 | 0.32 |
| Proportion of HIV positive people aged 15-29 on ART in 2017 | 0.49 | 0.57 | 0.57 | 0.57 | 0.57 | 0.57 | 0.57 | 0.57 | 0.57 | 0.57 | 0.57 | 0.57 | 0.57 | 0.57 | 0.57 | 0.57 |
| Proportion of HIV positive people aged 30-49 on ART in 2017 | 0.74 | 0.70 | 0.70 | 0.70 | 0.70 | 0.70 | 0.70 | 0.70 | 0.70 | 0.70 | 0.70 | 0.70 | 0.70 | 0.70 | 0.70 | 0.70 |
| Proportion of HIV positive people aged 50+ on ART in 2017 | 0.86 | 0.79 | 0.79 | 0.79 | 0.79 | 0.79 | 0.79 | 0.79 | 0.79 | 0.79 | 0.79 | 0.79 | 0.79 | 0.80 | 0.79 | 0.79 |
| Proportion of HIV positive men on ART in 2017 | 0.63 | 0.63 | 0.63 | 0.63 | 0.63 | 0.63 | 0.63 | 0.63 | 0.63 | 0.63 | 0.63 | 0.63 | 0.63 | 0.63 | 0.63 | 0.63 |
| Proportion of HIV positive women on ART in 2017 | 0.73 | 0.73 | 0.73 | 0.73 | 0.73 | 0.73 | 0.73 | 0.73 | 0.73 | 0.73 | 0.73 | 0.73 | 0.73 | 0.73 | 0.73 | 0.73 |
| Annual incidence of TB per 100,000 population in 2011 | 1433 (1107-1803) | 1194 | 1240 | 1228 | 1157 | 1193 | 1150 | 1175 | 1201 | 1179 | 1198 | 1242 | 1156 | 1256 | 1196 | 1161 |
| Annual incidence of TB per 100,000 population in 2018 | 658 (472-874) | 631 | 660 | 636 | 616 | 626 | 600 | 635 | 628 | 630 | 632 | 660 | 612 | 655 | 630 | 614 |
| Proportion of incident TB that is in HIV positive people in 2018 | 0.58 | 0.55 | 0.55 | 0.55 | 0.55 | 0.55 | 0.54 | 0.56 | 0.55 | 0.55 | 0.55 | 0.55 | 0.54 | 0.55 | 0.55 | 0.55 |
| Proportion of incident TB that is MDR in 2012 | 0.029 | 0.027 | 0.028 | 0.027 | 0.03 | 0.027 | 0.027 | 0.027 | 0.027 | 0.027 | 0.027 | 0.027 | 0.027 | 0.027 | 0.027 | 0.027 |
| Proportion of incident TB that is MDR in 2018 | 0.031 | 0.033 | 0.034 | 0.034 | 0.03 | 0.033 | 0.033 | 0.033 | 0.034 | 0.034 | 0.033 | 0.033 | 0.033 | 0.033 | 0.033 | 0.033 |
| Annual HIV- TB mortality rate per 100,000 population in 2018 | 47 (34-63) | 57 | 59 | 58 | 56 | 56 | 55 | 57 | 57 | 56 | 57 | 59 | 56 | 58 | 57 | 56 |

| | | | | | | | | | | | | | | | | |
|---|---|---|---|---|---|---|---|---|---|---|---|---|---|---|---|---|
| Annual HIV positive TB mortality rate per 100,000 population in 2016 | 92 (66-122) | 101 | 106 | 102 | 97 | 100 | 96 | 102 | 100 | 101 | 101 | 106 | 97 | 107 | 100 | 98 |
| Proportion of people starting TB treatment who are HIV positive in 2018 | 0.58 | 0.55 | 0.55 | 0.54 | 0.54 | 0.55 | 0.54 | 0.55 | 0.54 | 0.55 | 0.55 | 0.55 | 0.54 | 0.55 | 0.55 | 0.54 |
| Ratio of cases starting treatment to estimated incidence in 2000 | 57% (40-89) | 0.49 | 0.49 | 0.48 | 0.49 | 0.49 | 0.49 | 0.49 | 0.49 | 0.49 | 0.49 | 0.49 | 0.49 | 0.48 | 0.49 | 0.49 |
| Ratio of cases starting treatment to estimated incidence in 2018 | 76% (57-110) | 0.78 | 0.78 | 0.78 | 0.78 | 0.78 | 0.78 | 0.78 | 0.78 | 0.78 | 0.78 | 0.78 | 0.78 | 0.78 | 0.78 | 0.78 |
| Proportion starting treatment in 2017 who complete treatment, DS TB | 0.78 | 0.78 | 0.78 | 0.78 | 0.79 | 0.79 | 0.78 | 0.78 | 0.78 | 0.78 | 0.78 | 0.78 | 0.79 | 0.78 | 0.79 | 0.78 |
| Proportion starting treatment in 2017 who complete treatment, MDR TB | 0.54 | 0.54 | 0.54 | 0.54 | 0.54 | 0.55 | 0.53 | 0.52 | 0.54 | 0.54 | 0.53 | 0.54 | 0.53 | 0.54 | 0.54 | 0.54 |
| Proportion starting treatment in 2017 who die while on treatment, DS TB | 0.11 | 0.11 | 0.11 | 0.11 | 0.11 | 0.11 | 0.11 | 0.11 | 0.11 | 0.11 | 0.11 | 0.11 | 0.11 | 0.11 | 0.11 | 0.11 |
| Proportion starting treatment in 2017 who die while on treatment, MDR TB | 0.23 | 0.22 | 0.21 | 0.21 | 0.22 | 0.21 | 0.22 | 0.22 | 0.21 | 0.22 | 0.22 | 0.22 | 0.21 | 0.21 | 0.21 | 0.22 |
| Proportion starting treatment in 2017 who dropped out of treatment, DS TB | 0.11 | 0.11 | 0.11 | 0.11 | 0.10 | 0.10 | 0.11 | 0.11 | 0.11 | 0.11 | 0.10 | 0.11 | 0.10 | 0.11 | 0.10 | 0.10 |
| Proportion starting treatment in 2017 who dropped out of treatment, MDR TB | 0.23 | 0.23 | 0.24 | 0.23 | 0.22 | 0.22 | 0.23 | 0.24 | 0.23 | 0.24 | 0.24 | 0.23 | 0.24 | 0.24 | 0.24 | 0.23 |
| Increased prevalence of TB in clinic attendees, compared to the general population | 1.86 | 1.83 | 1.83 | 1.83 | 1.83 | 1.83 | 1.82 | 1.83 | 1.29* | 3.09* | 1.84 | 1.83 | 1.84 | 1.81 | 1.83 | 1.83 |
| Proportion of incident TB that results from transmission between household members, in 2018 | 8-19% | 0.12 | 0.12 | 0.12 | 0.17* | 0.07* | 0.12 | 0.12 | 0.12 | 0.12 | 0.12 | 0.12 | 0.12 | 0.12 | 0.12 | 0.12 |
| Relative change in HIV prevalence in men between 2020 and 2030‡ | -0.16 | -0.16 | -0.16 | -0.16 | -0.16 | -0.16 | -0.16 | -0.16 | -0.16 | -0.16 | -0.16 | -0.16 | -0.16 | -0.16 | -0.26* | 0.038* |
| Relative change in HIV prevalence in women between 2020 and 2030‡ | -0.057 | -0.058 | -0.058 | -0.058 | -0.059 | -0.058 | -0.058 | -0.059 | -0.058 | -0.059 | -0.058 | -0.058 | -0.058 | -0.058 | -0.17* | 0.067* |

**Table S12. Model fit to fitting targets, in the best estimate scenario and sensitivity analysis scenarios.** 'Low' and 'high' refer to changes that decrease and increase the proportion of disease that results from transmission in clinics respectively. *Indicates fitting outputs where the target value was changed in the sensitivity analysis. ┼Indicates outputs where the value could change in the intervention scenarios. Figures shown are for the baseline scenario

#### 3.3 Results by uncertainty analysis scenario

##### 3.3.1 Proportion of disease from transmission in clinics

Figure S3 shows the proportion of disease that resulted from transmission in clinics in the study population in 2019, by scenario and by population group. The sources of uncertainty in model input parameters that had the largest effect on model estimates were the amount of contact time that occurred in clinics, the prevalence of TB in clinic attendees compared to the general population, and ventilation levels in clinics relative to in other settings. The proportion of disease that results from transmission in households, and the rate at which individuals switched between high and low clinic visiting groups, had little effect on model estimates.

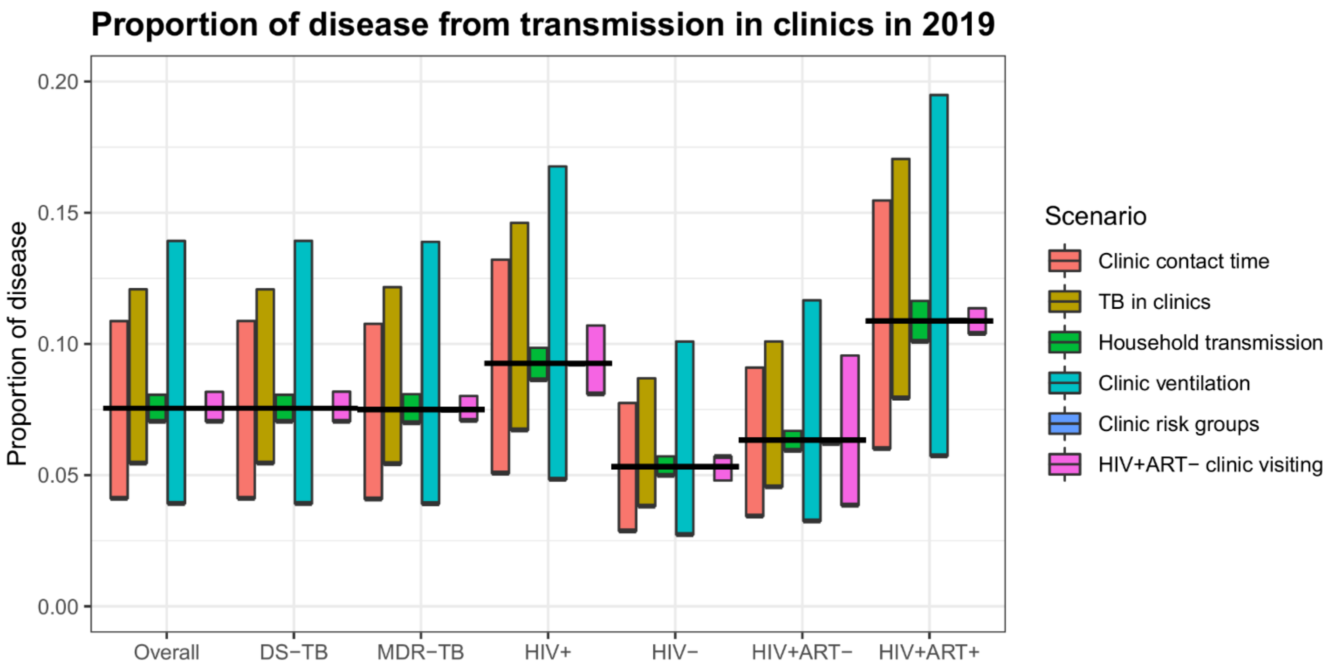

**Figure S3. The estimated proportion of disease that resulted from transmission in clinics in the study population in 2019, by scenario and by population group.** Horizontal black lines show the estimates from the 'best estimate' scenario. See section 'Uncertainty analyses' for a description of the scenarios. The 'Clinic risk groups' uncertainty analysis had little effect on the results, and therefore the bar is mostly hidden under the horizontal black lines.

##### 3.3.2 Intervention impact

Figures S4-S7 show the estimated reductions in TB cases and TB deaths, overall and MDR-TB, in the study population in 2021-2030, by intervention and scenario.

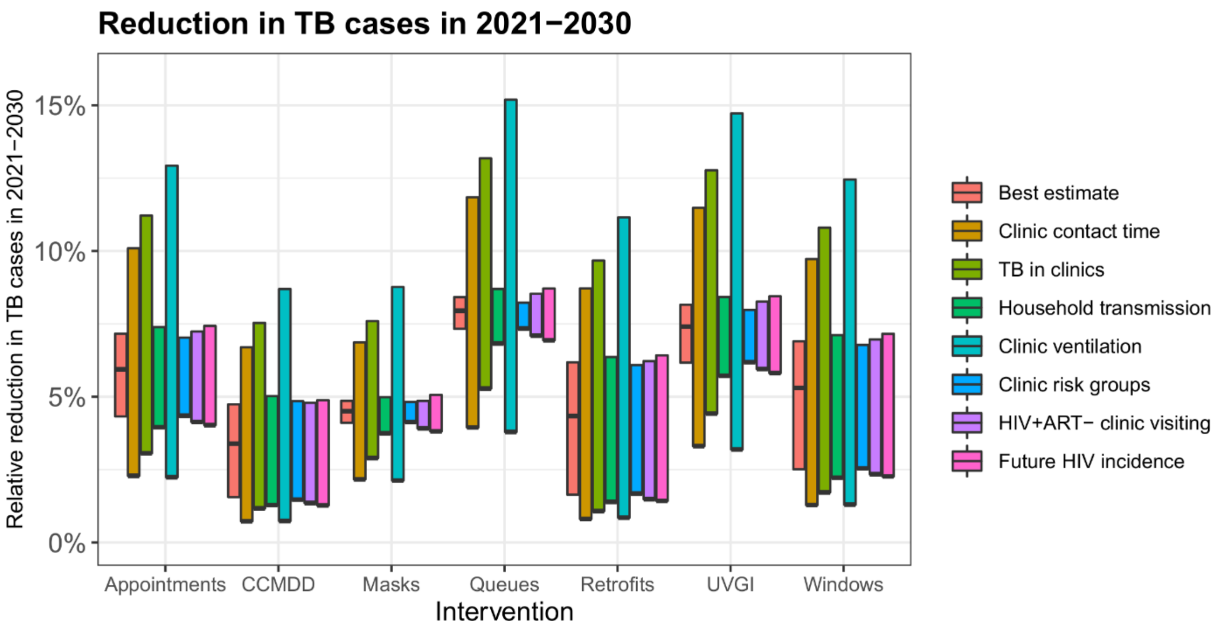

**Figure S4. The estimated reduction in TB cases in the study population in 2021-2030 resulting from the proposed infection prevention and control interventions, by scenario.**

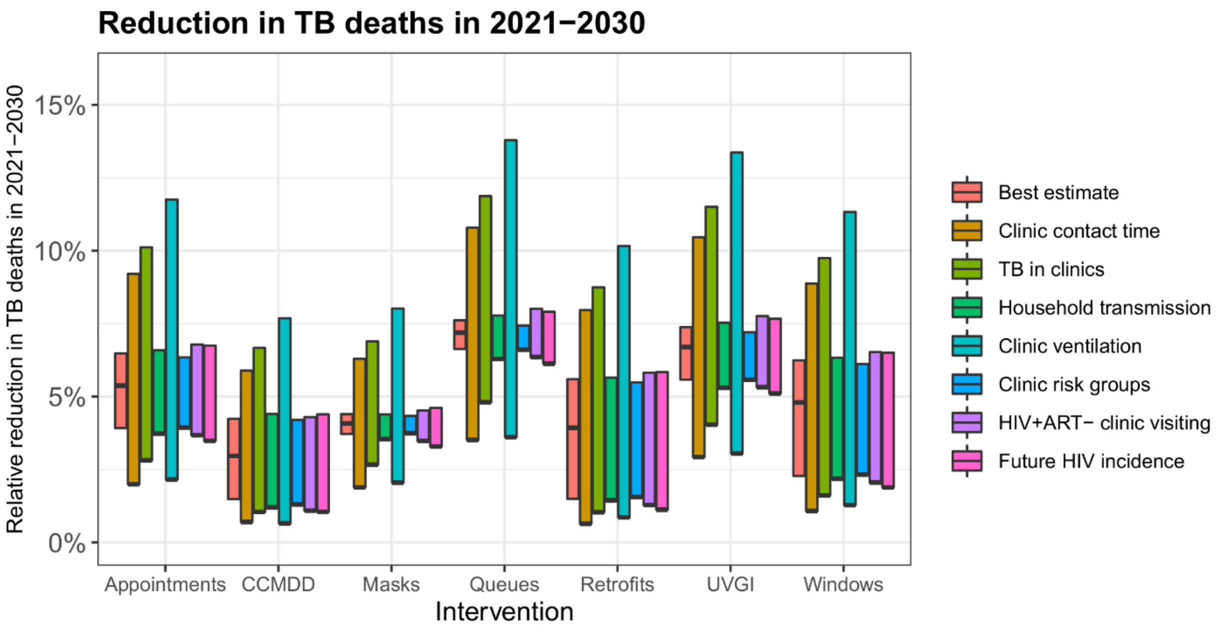

**Figure S5. The estimated reduction in TB deaths in the study population in 2021-2030 resulting from the proposed infection prevention and control interventions, by scenario.**

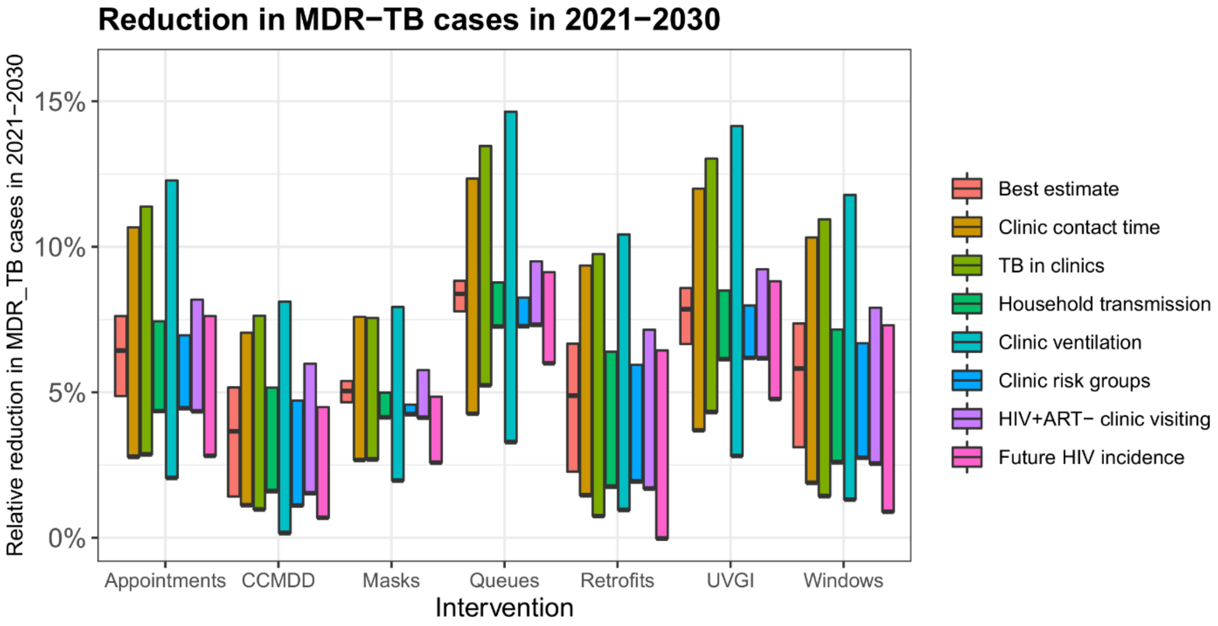

**Figure S6. The estimated reduction in MDR-TB cases in the study population in 2021-2030 resulting from the proposed infection control interventions, by scenario.**

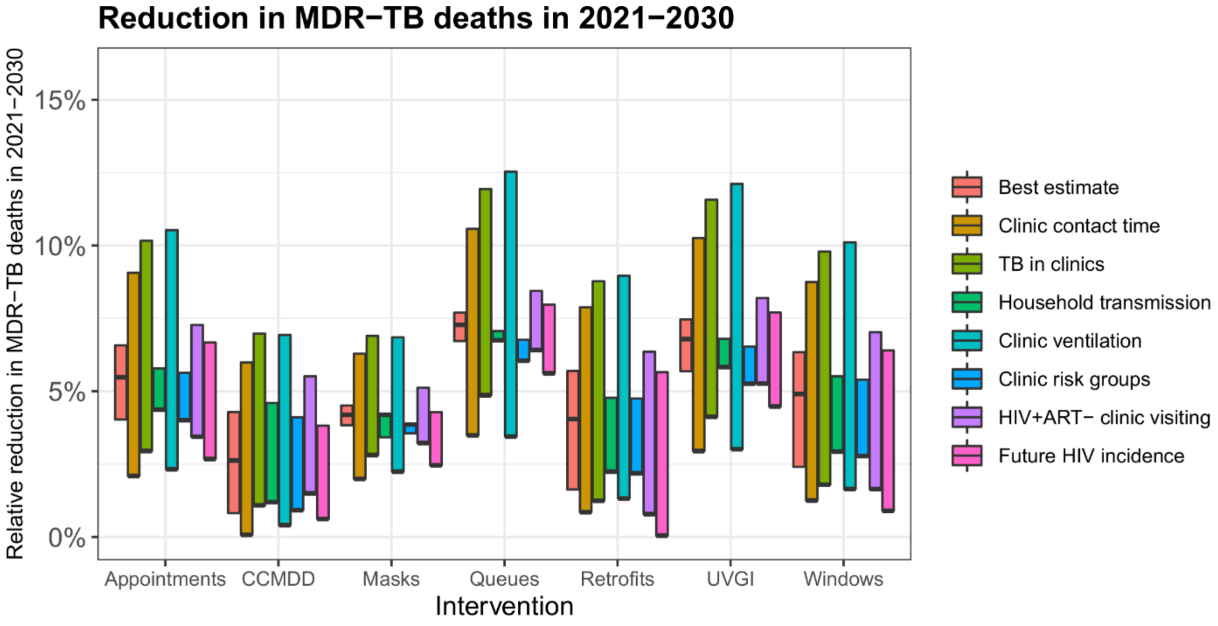

**Figure S7. The estimated reduction in MDR-TB deaths in the study population in 2021-2030 resulting from the proposed infection control interventions, by scenario.**

### 4 Proportion of disease from transmission in clinics that is in clinic staff

#### 4.1 Methods

In the mathematical model, we only consider transmission to adult clinic attendees (patients, and people attending with or on the behalf of patients). Clinic staff are also at risk of infection in clinics however, and here we use a simple calculation to obtain a rough estimate of the proportion of tuberculosis in adults resulting from transmission in clinics that is in clinic staff.

The proportion can be estimated using the following equation:

$$p = s(s + c(1 - s)(r - 1)^{-1})^{-1}$$

Where:

- $p$ is the proportion of all disease resulting from transmission in clinics that is in clinic staff
- $s$ is the proportion of the population who are clinic staff
- $c$ is the proportion of disease that results from transmission in clinics in the general population
- $r$ is the relative rate of TB in clinic staff compared to the general population

Assuming that all clinic staff who are at elevated risk of infection from transmission in clinics have the same exposure to TB outside the clinic as the general population, and that all excess TB in clinic staff results from transmission in clinics.

Two clinics serve the study population. The clinics have a total of 59 staff who are considered to be at elevated risk of infection from transmission in clinics, with the rest being situated outside the majority of the time (e.g. security guards), or spending little time in public areas when patients are present. The adult total population of the study communities was 33,288. This means that $s = 59 / 33288$.

The results of this work indicate that 7.1% of disease in adults in the general population results from transmission in the clinic, with a plausible range of 4.0-14.2%.

No data were available on excess tuberculosis risk in clinic staff in our study setting. A recent systematic review of TB incidence in healthcare workers estimated that the ratio of the rate of TB in healthcare workers compared to the general population in high TB burden settings was 4.32 (95% CI 2.36-7.91[22]).

To generating a best estimate for *p*, we took the best estimates for all three parameters. To generate a 95% range, we generated 10,000 bootstrap samples, sampling *c* from uniform(0.04,0.142), and *r* from a split normal distribution with mean 4.32 and 95% CI 2.36-7.91. The 95% range was calculated as the 0.025$^{th}$ and 0.975$^{th}$ percentiles.

#### 4.2 Results

We estimate that in the study community, an average of 7.1% (95% plausible range 2.3-16.7%) of all disease in adults resulting from transmission in clinics occurs in clinic staff.

### 5 References

1. McCreesh N, Dlamini V, Edwards A, et al. Impact of social distancing regulations and epidemic risk perception on social contact and SARS-CoV-2 transmission potential in rural South Africa: analysis of repeated cross-sectional surveys. *medRxiv* 2020;2020.12.01.20241877 doi: https://doi.org/10.1101/2020.12.01.20241877
2. Middelkoop K, Bekker L-G, Morrow C, et al. Decreasing household contribution to TB transmission with age: a retrospective geographic analysis of young people in a South African township. *BMC Infectious Diseases* 2014;14(1):221. doi: 10.1186/1471-2334-14-221
3. Govender I, Karat AS, Baisley K, et al. Prevalence of M. tuberculosis in sputum among clinic attendees compared with the surrounding community in rural South Africa: implications for finding the missing millions. 51st Union World Conference on Lung Health, 2020.
4. Lygizos M, Shenoi SV, Brooks RP, et al. Natural ventilation reduces high TB transmission risk in traditional homes in rural KwaZulu-Natal, South Africa. *BMC Infectious Diseases* 2013;13(1):300. doi: 10.1186/1471-2334-13-300
5. Beckwith P, Deol A, McCreesh N, et al. Direct estimates of absolute ventilation in primary health care clinics in South Africa.
6. Taylor JG, Yates TA, Mthethwa M, et al. Measuring ventilation and modelling M. tuberculosis transmission in indoor congregate settings, rural KwaZulu-Natal. *Int J Tuberc Lung Dis* 2016;20(9):1155-61. doi: 10.5588/ijtld.16.0085 [published Online First: 2016/08/12]
7. Organization WH. Rapid communication: key changes to treatment of multidrug-and rifampicin-resistant tuberculosis (MDR/RR-TB): World Health Organization, 2018.
8. Aurum Institute. Managing TB - In A New Era Of Diagnostics, 2016.
9. Glynn JR, Murray J, Bester A, et al. High rates of recurrence in HIV-infected and HIV-uninfected patients with tuberculosis. *The Journal of infectious diseases* 2010;201(5):704-11.
10. Verver S, Warren RM, Beyers N, et al. Rate of Reinfection Tuberculosis after Successful Treatment Is Higher than Rate of New Tuberculosis. *American Journal of Respiratory and Critical Care Medicine* 2005;171(12):1430-35. doi: 10.1164/rccm.200409-1200OC
11. World Health Organization. Global tuberculosis report 2019. Geneva, Switzerland: World Health Organization; 2019, 2019.
12. Yates TA. Mycobacterium tuberculosis infection in Southern Africa–exploring patterns, locating transmission. UCL (University College London), 2016.
13. Middelkoop K, Bekker L-G, Myer L, et al. Rates of tuberculosis transmission to children and adolescents in a community with a high prevalence of HIV infection among adults. 2008;47(3):349-55.
14. McCreesh N, White RG. An explanation for the low proportion of tuberculosis that results from transmission between household and known social contacts. *Scientific reports* 2018;8(1):5382.

15. Houben RMGJ, Lalli M, Sumner T, et al. TIME Impact – a new user-friendly tuberculosis (TB) model to inform TB policy decisions. *BMC Medicine* 2016;14(1):56. doi: 10.1186/s12916-016-0608-4
16. Dowdy DW, Chaisson RE. The persistence of tuberculosis in the age of DOTS: reassessing the effect of case detection. *Bulletin of the World Health Organization* 2009;87(4):296-304. doi: 10.2471/BLT.08.054510
17. Vandormael A, Akullian A, Siedner M, et al. Declines in HIV incidence among men and women in a South African population-based cohort. 2019;10(1):1-10.
18. Johnson LF, Dorrington RE, Moolla H. Progress towards the 2020 targets for HIV diagnosis and antiretroviral treatment in South Africa. *Southern African journal of HIV medicine* 2017;18(1)
19. https://thembisa.org/ [accessed 24/8/2020.
20. Diaconu K, Karat AS, Bozzani F, et al. Systems interventions for improving TB infection prevention and control in South African primary care facilities
21. McCreesh N, Karat AS, Baisley K, et al. Modelling the effect of infection prevention and control measures on rate of Mycobacterium tuberculosis transmission to clinic attendees in primary health clinics in South Africa. *medRxiv* 2021;2021.07.26.21260835 doi: https://doi.org/10.1101/2021.07.26.21260835
22. World Health Organization. WHO guidelines on tuberculosis infection prevention and control: 2019 update: World Health Organization 2019.
23. Chartier Y, Pessoa-Silva C. Natural ventilation for infection control in health-care settings: World Health Organization 2009.
24. Mphaphlele M, Dharmadhikari AS, Jensen PA, et al. Institutional tuberculosis transmission. Controlled trial of upper room ultraviolet air disinfection: A basis for new dosing guidelines. *American Journal of Respiratory and Critical Care Medicine* 2015;192(4):477-84. doi: 10.1164/rccm.201501-0060OC
25. Dharmadhikari AS, Mphahlele M, Stoltz A, et al. Surgical face masks worn by patients with multidrug-resistant tuberculosis: impact on infectivity of air on a hospital ward. 2012;185(10):1104-09.
26. MacIntyre CR, Chughtai AAJB. Facemasks for the prevention of infection in healthcare and community settings. 2015;350:h694.
27. Health Systems Trust. The CCMDD story, 2019.
28. HST Indicator Tool [Available from: https://indicators.hst.org.za/ accessed 7/4/2020 2020.
29. Karat AS, McCreesh N, Baisley K, et al. Waiting times, occupancy density, and patient flow in South African primary health clinics: implications for infection prevention and control. *MedRxiv* 2021;2021.07.21.21260806 doi: https://doi.org/10.1101/2021.07.21.21260806
30. Escombe AR, Ticona E, Chávez-Pérez V, et al. Improving natural ventilation in hospital waiting and consulting rooms to reduce nosocomial tuberculosis transmission risk in a low resource setting. *BMC infectious diseases* 2019;19(1):88.
31. Menzies NA, Cohen T, Lin H-H, et al. Population health impact and cost-effectiveness of tuberculosis diagnosis with Xpert MTB/RIF: a dynamic simulation and economic evaluation. *PloS Med* 2012;9(11):e1001347.
32. Ragonnet R, Flegg JA, Brilleman SL, et al. Revisiting the Natural History of Pulmonary Tuberculosis: A Bayesian Estimation of Natural Recovery and Mortality Rates. *Clinical Infectious Diseases* 2020 doi: 10.1093/cid/ciaa602
33. Dheda K, Lampe FC, Johnson MA, et al. Outcome of HIV-Associated Tuberculosis in the Era of Highly Active Antiretroviral Therapy. *The Journal of Infectious Diseases* 2004;190(9):1670-76. doi: 10.1086/424676
34. Lawn SD, Kranzer K, Wood RJCicm. Antiretroviral therapy for control of the HIV-associated tuberculosis epidemic in resource-limited settings. 2009;30(4):685-99.
35. Kasaie P, Andrews JR, Kelton WD, et al. Timing of Tuberculosis Transmission and the Impact of Household Contact Tracing: An Agent-Based Simulation Model. *American Journal of*

*Respiratory and Critical Care Medicine* 2014;189(7):845-52. doi: 10.1164/rccm.201310-1846OC
36. Lawn SD, Myer L, Edwards D, et al. Short-term and long-term risk of tuberculosis associated with CD4 cell recovery during antiretroviral therapy in South Africa. 2009;23(13):1717.
37. Corbett EL, Watt CJ, Walker N, et al. The growing burden of tuberculosis: global trends and interactions with the HIV epidemic. *Archives of Internal Medicine* 2003;163(9):1009-21.
38. Ismail NA, Mvusi L, Nanoo A, et al. Prevalence of drug-resistant tuberculosis and imputed burden in South Africa: a national and sub-national cross-sectional survey. 2018;18(7):779-87.
39. Mandela N. Nelson Mandela/HSRC study of HIV/AIDS: South African national HIV prevalence, behavioural risks and mass media: household survey 2002: HSRC Press 2002.
40. Mossong J, Grapsa E, Tanser F, et al. Modelling HIV incidence and survival from age-specific seroprevalence after antiretroviral treatment scale-up in rural South Africa. *AIDS (London, England)* 2013;27(15):2471-79. doi: 10.1097/01.aids.0000432475.14992.da
41. Brinkhof MW, Boulle A, Weigel R, et al. Mortality of HIV-infected patients starting antiretroviral therapy in sub-Saharan Africa: comparison with HIV-unrelated mortality. 2009;6(4)
42. Granich R, Gupta S, Hersh B, et al. Trends in AIDS Deaths, New Infections and ART Coverage in the Top 30 Countries with the Highest AIDS Mortality Burden; 1990-2013. *PLoS One* 2015;10(7):e0131353-e53. doi: 10.1371/journal.pone.0131353
43. Statistics South Africa. Mid-year population estimates 2019. Pretoria, South Africa, 2019.
44. Statistics South Africa. Mid-year population estimates 2018. Pretoria, South Africa, 2018.

### 6 Acknowledgements

The extended *Umoya omuhle* team, institutions, and roles (listed alphabetically by surname):

| Name | Institution/s | Role |
|---|---|---|
| Siphokazi Adonisi | UCT | Research Assistant |
| Kathy Baisley | LSHTM; AHRI | Co-investigator |
| Peter Beckwith | LSHTM; UCT | Research fellow |
| Fiammetta Bozzani | LSHTM | Co-investigator |
| Amy Burdzik | UCT | Occupational health |
| Adrienne Burrough | LSHTM | Project Manager |
| Nkosingiphile Buthelezi | AHRI | Research Assistant |
| Xolile Buthelezi | AHRI | Diagnostic Lab Manager |
| Ruvimbo Chigwanda | UCT | Administration |
| Christopher Colvin | UCT | Co-investigator |
| PIP CRAs | AHRI | Clinic research Assistants |
| Njabulo Dayi | AHRI | Research Data Manager |
| Arminder Deol | LSHTM | Mathematical modeller |
| Karina Diaconu | QMU | Co-investigator |
| Siphephelo Dlamini | AHRI | Nursing Manager |
| Yutu Dlamini | AHRI | Research Assistant |
| Raveshni Durgiah | AHRI | Grants office |
| Anita Edwards | AHRI | Head: Scientific Support |
| Jennifer Falconer | QMU | Research Assistant |
| Kitty Flynn | QMU | Administrator |
| Patrick Gabela | AHRI | Clinical Research Data Coordinator |
| Dickman Gareta | AHRI | Head: Research Data Management |
| Awethu Gawulekapa | UCT | Research Assistant |
| Harriet Gliddon | AHRI; UCL | Research Assistant |
| Bavashni Govender | UKZN | Administration |
| Indira Govender | LSHTM; AHRI | Co-investigator |
| Alison Grant | LSHTM; AHRI | Principal investigator |
| Meghann Gregg | LSE | Research fellow |
| Emmerencia Gumede | AHRI | Research Assistant |
| Sashin Harilall | AHRI | Grants office |
| Kobus Herbst | AHRI | Chief Information Officer |
| Tamia Jansen | UCT | Research Assistant |
| Seonaid Kabiah | UCT | Research Assistant |
| Idriss Kallon | UCT | Post-doctoral researcher |
| Aaron Karat | LSHTM | Co-investigator |
| Hannah Keal | AHRI | Communications |
| Suzanne Key | UCT | Occupational health |
| Zama Khanyile | UKZN | Research Assistant |
| Mandla Khoza | AHRI | Clinic Research Assistant |
| Nozi Khumalo | AHRI | Systems Engineer |

| Name | Institution/s | Role |
|---|---|---|
| Zilethile Khumalo | AHRI | Research Assistant |
| Karina Kielmann | QMU | Co-principal investigator |
| Nondumiso Kumalo | AHRI | Clinic Research Assistant |
| Richard Lessells | AHRI | Epidemiologist |
| Nokuthula Lushaba (deceased) | UKZN | Administration |
| Sithembiso Luthuli | AHRI | Research Assistant |
| Sinethemba Mabuyakhulu | AHRI | Clinic Research Assistant |
| Hayley MacGregor | IDS | Co-investigator |
| Nonhlanhla Madlopha | AHRI | Research Assistant |
| Aphiwe Makalima | UCT | Administration |
| Tacha Malaza | AHRI | PIP CRA |
| Sifundesihle Malembe | AHRI | Research Assistant |
| Godfrey Manuel | UCT | Transport |
| Nonhlanhla Maphumulo | UKZN | Administration |
| Precious Mathenjwa | UCT | Research Assistant |
| Sanele Mbuyazi | AHRI | PIP CRA |
| Nicky McCreesh | LSHTM | Co-investigator |
| Claire McLellan | QMU | Administrator |
| Simphiwe Mdluli | AHRI | PIP CRA |
| Thabile Mkhize | AHRI | Transport |
| Duduzile Mkhwanazi | AHRI | Research Assistant |
| Zinhle Mkhwanazi | AHRI | Research Assistant |
| Zodwa Mkhwanazi | AHRI | Research Assistant |
| Anathi Mngxekeza | UCT | Research Assistant |
| Tshwaraganang Modise | AHRI | Research Data |
| Sashen Moodley | AHRI | Microbiology Laboratory Supervisor |
| Samantha Moyo | UCT | Research Assistant |
| Silindile Mthembu | AHRI | Clinic Research Assistant |
| Nozipho Mthethwa | AHRI | Research Assistant |
| Siphesihle Mthethwa | AHRI | Procurement Coordinator |
| Sphiwe Mthethwa | AHRI | Research Assistant |
| Sanele Mthiyane | AHRI | Research Assistant |
| Vanisha Munsamy | AHRI | Grants office |
| Sinead Murphy | UCT | Research Assistant |
| Thomas Murray | AHRI | Research assistant |
| Senzile Myeni | AHRI | PIP CRA |
| Tevania Naidoo | AHRI | Procurement |
| Nompilo Ndlela | AHRI | Research Assistant |
| Zama Ndlela | AHRI | PIP CRA |
| Thandekile Nene | AHRI | Research Assistant |
| Phumla Ngcobo | AHRI | Communications |
| Nzuzo Ntombela | AHRI | Research Data Systems Service Manager |
| Sabelo Ntuli | AHRI | GIS Coordinator |
| Nompumulelo Nyawo | AHRI | Human resources |

| Name | Institution/s | Role |
|---|---|---|
| Phumzile Nywagi | UCT | Research Assistant |
| Stephen Olivier | AHRI | Statistician |
| Justin Parkhurst | LSE | Co-investigator |
| Alex Pym | AHRI | Co-investigator |
| Yolanda Qeja | UCT | Research Assistant |
| Anand Ramnanan (deceased) | AHRI | Procurement |
| Sharmila Rugbeer | UKZN | Administration |
| Janet Seeley | LSHTM | Co-investigator |
| Aruna Sevakram | AHRI | Scientific support |
| Sizwe Sikhakane | AHRI | Transport |
| Zizile Sikhosana | AHRI | Somkhele Laboratory Supervisor |
| Theresa Smit | AHRI | Head: Diagnostic Research |
| Thandeka Smith | UKZN | Research Assistant |
| Naomi Stewart | LSHTM | Communications |
| Alison Swartz | UCT | Co-investigator |
| Amy Thomas | LSHTM | Communications |
| Siphosethu Titise | UCT | Research Assistant |
| Anna Vassall | LSHTM | Co-investigator |
| Marlise Venter | AHRI | Facilities Administrator |
| Anna Voce | UKZN | Co-investigator |
| Richard White | LSHTM | Co-investigator |
| Tom Yates | Imperial | Co-investigator |
| Precious Zulu | AHRI | Administration |
| Gimenne Zwama | QMU | Research Fellow |

AHRI: Africa Health Research Institute; IDS: Institute of Development Studies; LSE: London School of Economics and Political Science; LSHTM: London School of Hygiene & Tropical Medicine; QMU: Queen Margaret University; UCT: University of Cape Town; UKZN: University of KwaZulu-Natal;
